# A multi-ancestry genome-wide meta-analysis, fine-mapping, and gene prioritization approach to characterize the genetic architecture of adiponectin

**DOI:** 10.1101/2023.05.02.23289402

**Authors:** Vishal Sarsani, Sarah M. Brotman, Yin Xianyong, Lillian Fernandes Silva, Markku Laakso, Cassandra N. Spracklen

## Abstract

Previous genome-wide association studies (GWAS) for adiponectin, a complex trait linked to type 2 diabetes and obesity, identified >20 associated loci. However, most loci were identified in populations of European ancestry, and many of the target genes underlying the associations remain unknown. We conducted a multi-ancestry adiponectin GWAS meta-analysis in ≤46,434 individuals from the METSIM cohort and the ADIPOGen and AGEN consortiums. We combined study-specific association summary statistics using a fixed-effects, inverse variance-weighted approach. We identified 22 loci associated with adiponectin (*P* < 5×10^−8^), including 15 known and 7 previously unreported loci. Among individuals of European ancestry, GCTA-COJO identified 14 additional distinct signals at the *ADIPOQ*, *CDH13*, *HCAR1*, and *ZNF664* loci. Leveraging the multi-ancestry data, FINEMAP + SuSiE identified 46 causal variants (PP>0.9), which also exhibited potential pleiotropy for cardiometabolic traits. To prioritize target genes at associated loci, we propose a combinatorial likelihood scoring formalism (“GPScore”) based on measures derived from 11 gene prioritization strategies and the physical distance to the transcription start site. With “GPScore”, we prioritize the 30 most probable target genes underlying the adiponectin-associated variants in the multi-ancestry analysis, including well-known causal genes (e.g., *ADIPOQ*, *CDH13*) and novel genes (e.g., *CSF1*, *RGS17*). Functional association networks revealed complex interactions of prioritized genes, their functionally connected genes, and their underlying pathways centered around insulin and adiponectin signaling, indicating an essential role in regulating energy balance in the body, inflammation, coagulation, fibrinolysis, insulin resistance, and diabetes. Overall, our analyses identify and characterize adiponectin association signals and inform experimental interrogation of target genes for adiponectin.

## Introduction

Adiponectin, an abundant adipocytokine secreted almost exclusively by adipocytes, is crucial in the interconnection between adiposity, insulin resistance, and inflammation ^1–3^. Previous genetic studies have identified more than 20 loci harboring variants associated with serum levels of adiponectin, including *ADIPOQ* and *CDH13* ^4–7^. Genetic analyses have also indicated shared allelic architecture between adiponectin, type 2 diabetes, and other metabolic traits (e.g., BMI, WHR) ^4, 7, 8^. Due to its diverse physiological functions in glucose and lipid metabolism, inflammation, and oxidative stress across metabolic and cardiovascular tissues, adiponectin could be a possible therapeutic target for metabolic syndrome, diabetes, and coronary disease ^9^. However, to move established loci toward effective clinical and therapeutic targets, the functional variant(s), target/effector gene(s), and the mechanistic direction(s) of effect need to be identified; for most loci identified to date, this information remains largely unknown.

Multiple methods have been proposed to prioritize target genes underlying genome-wide association study (GWAS) signals using expression, functional genomics, and network data ^10–12^. However, individual approaches often have conflicting findings, making it difficult to interpret or prioritize candidate target genes. Attempts to incorporate integrative and complementary gene prioritization approaches to identify disease-risk genes have been somewhat successful ^13, 14^. However, the primary challenge in using prioritization approaches for a complex trait or disease is that the lack of customizability to focus on the integration of the data that is most relevant (e.g., tissue specificity) to the disease or trait of interest. This may result in a researcher choosing results in an ad hoc or post-hoc manner. A researcher may also want to maximize the likelihood of selecting a suitable target gene for experimental follow-up, and considering support from multiple approaches will make the selection(s) more robust.

Leveraging summary statistics from diverse genetic studies, we conducted a multi-ancestry, genome-wide meta-analysis for adiponectin in up to 46,434 individuals from the Metabolic Syndrome in Men (METSIM) cohort and the ADIPOGen and AGEN consortiums ^4, 5, 15^. Our primary objectives were to: 1) discover previously unreported loci associated with plasma adiponectin levels; 2) narrow putative causal variants underlying the association signals; 3) prioritize target genes systematically by using our proposed “GPScore” approach based on evidence derived from 11 gene prioritization strategies and physical distance to transcription start sites; and 4) perform functional profiling of target genes and their underlying pathways.

## Materials and methods

### Ethics statement

The research protocol for all studies was reviewed and approved by the institutional ethics review committees at the involved institutions. Written informed consent was obtained from all study participants.

### Study Design and Participants

Our meta-analysis included summary statistics from 16 studies of European-ancestry individuals from the ADIPOGen consortium (n = 29,347 from the discovery phase) ^4^, four studies with individuals of East Asian-ancestry from the AGEN consortium (n=7,825) ^5^, and the METSIM study (European-ancestry; n=9,262) ^15^. While most of our analyses were performed in both European-only and all ancestries combined (“multi-ancestry”), we consider the multi-ancestry analyses to be our primary results. A detailed description of participant characteristics, genotype and phenotype information, quality control, and imputation can be found in Table S1.

### Genome-wide meta-analyses

We performed the European-ancestry and multi-ancestry adiponectin meta-analyses using a fixed-effects inverse variance-weighted meta-analysis approach with the random-effects model (RE2) implemented in METASOFT ^16^, which corrects for population structure while allowing for examination of heterogeneity statistics. In the multi-ancestry meta-analysis, we also generated Bayes factors (BF) from MR-MEGA ^17^, which employs meta-regression to account for heterogeneity in allelic effects associated with ancestry. We defined the “lead” association signal at each locus to be the most significant variant (*P* < 5×10^−8^) within a 500 kb window. We considered association signals to be previously unreported, or “novel”, if they were located > 500 kb from a previously reported adiponectin signal. To identify distinct association signals at each locus identified in the European-ancestry analysis ^4–6, 15, 18^, we performed approximate conditional analyses using genome-wide joint conditional analysis (COJO) implemented in the Genome-wide Complex Traits Analysis (GCTA) software ^19^. Using data from 10,197 METSIM participants to calculate linkage disequilibrium (LD) and a collinearity threshold of (*R*^2^) = 0.9, we considered a variant to represent an additional, distinct signal at a locus if: 1) the variant achieved (*P* < 5×10^−8^) in the COJO analysis, and 2) the variant was located within +/-1 Mb from the original lead variant at that locus. We use “index variant to denote the most significant variant within each of the secondary association signals.

### Pleiotropic associations with phenotypes from CMDKP

We interrogated our lead and index variants with data from the Common Metabolic Diseases Knowledge Portal (www.cmdkp.org) to explore pleiotropic associations between the index variants from the multi-ancestry meta-analysis and other complex traits across five common disease areas (type 1 and type 2 diabetes, cardiovascular and cerebrovascular disease, and sleep disorders). We only considered traits/diseases that achieved *P*< 0.05 in the original analyses. For ease of visualization, we aggregated traits/diseases into 23 broad categories.

### Identifying candidate causal variants

We used several approaches to identify candidate causal variants at loci identified in both the multi-ancestry and European ancestry-only meta-analyses. Unless otherwise stated, we used data from 10,197 METSIM participants as the reference for LD calculations.

### Statistical fine-mapping

We performed statistical fine-mapping using FINEMAP ^20^ and SuSiE^21^. At each adiponectin-associated locus, we computed in-sample dosage LD using LDstore ^22^. We defined a fine-mapping region as the 3 Mb window (+/- 1.5 Mb) around each lead variant. We allowed up to 10 causal variants per window and extracted the posterior inclusion probabilities (PIP) of each variant using each method independently. The variants with a PIP > 0.90 in either of the fine-mapping methods, along with having LD *r*^2^ > 0.8 with the lead variant, are considered the final candidate causal variants.

### Causal variant annotation

The majority of the candidate causal variants reside in non-coding regions of the human genome. We used RegulomeDB ^23^ to annotate the candidate causal variants from fine-mapping with evidence of regulatory function(s) through functional genomic assays and computational approaches. RegulomeDB provides a score (range 1-7) indicating its potential to be functional in regulatory elements and the probability of confidence in the score for each variant. We interrogated fine-mapped candidate causal variants from the multi-ancestry fine-mapping using CAUSALdb ^24^, a database containing fine-mapping results from over 3,052 GWAS summary statistics. Whenever multiple variants mapped to the same trait across different studies, we meta-analyzed the different p-values together.

### Gene Prioritization

We created the Gene Priority Score (GPScore), which combines evidence from 11 gene prioritization strategies ^11, 21, 25–33^, along with the physical distance to the transcription start site(s) (TSS), to prioritize target genes underlying our adiponectin association signals. For all approaches, we considered all protein-coding genes within the +/- 1.5 Mb of the original lead variant from the multi- or European-ancestry meta-analyses. For prioritization solely based on gene expression ^21, 27–30^, we restricted our analysis to tissues most biologically-relevant to adiponectin (adipose subcutaneous, adipose visceral omentum, adrenal gland, artery aorta, artery coronary, artery tibial, heart atrial appendage, heart left ventricle, kidney cortex, liver, muscle-skeletal, thyroid, and whole blood). Unless otherwise stated, we used data from 10,197 METSIM participants as the reference for LD calculations.

#### Gene Prioritization Score

GPScore is a combinatorial likelihood score constructed by leveraging various gene prioritization strategies described below. The score is unbiased and only involves weighting factors to prioritize signals from a particular tissue. GPScore is defined as

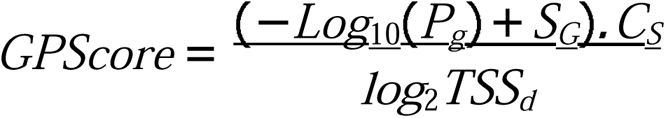

Where *P_G_* is a combined P-value for MAGMA ^25^, summary Mendelian randomization (SMR) ^28^, expression quantitative trait loci (eQTL) colocalization (LD-based approach), SNP-heritability enrichment ^29^ and Downstreamer ^32^ (described below), computed by the sum of z via Stoufferâs method^34^. The second term (*S_G_*) represents the combined scores of eQTL colocalization (coloc) ^35^, EMS ^30^, PoPS ^31^, EpiMap ^11^, cS2G ^33^ and Genehancer ^18^ scores. We normalized scores for EMS, PoPS, and Genehancer scores by scaling between 0 and 1. The third term (*C_S_*) is a score ranging from 0 to 1, representing the proportion of support for a particular protein-coding gene across all 11 gene prioritization strategies. Each strategy has a maximum support score of 1. For eQTL colocalizations (coloc), SNP-heritability enrichment, EMS score, and SMR, we assigned a score of one if the tissue source was adipose or cardiac-related tissue (given the trait of interest is adiponectin); else, we assigned a score of 0.8. The final term (*TSS_d_*) is the distance from the lead variant to the transcription start site of the gene/transcript, measured in base pairs.

One of the methodological issues in computing GPScore is that some of the gene prioritization strategies (i.e., coloc, EMS, SMR, GeneHancer, cs2G) report values for individual variants/position, whereas other strategies prioritize genes (i.e., MAGMA, Downstreamer, Epimap, PoPS, LDSC). We transformed variant/position level scores into gene-level scores using recommendations from Lehne et al. ^36^. For expression-based enrichments, a variant/position may have a statistic corresponding to a tissue (e.g., colocalization of a variant in adipose tissue). Using the average or highest quartile of a statistic may result in the inability to assign tissue information after the final transformation. Because preserving tissue information in the variant scores is essential for weighing support (*C_S_*), we have opted to use either the lowest p-value (MAGMA, SMR, eQTL colocalization LD-based approach, SNP-heritability enrichment, Downstreamer) or the maximum score (coloc, EMS, PoPS, EpiMap, Genehancer) per gene. Alternatively, in situations where tissue information is less critical, the average or highest quartile of statistics could be used.

#### MAGMA gene analysis

To quantify the degree of association of each gene with adiponectin while incorporating LD structure between variants, we used MAGMA v1.10^25^ to perform gene-set analyses and obtain gene P-values from the multiple linear principal components regression F-test. The null hypothesis of the F-test is that the gene has no effect on the phenotype, conditional on all covariates. The variant-wise mean model was used, and variants were assigned to one of the 18,383 (GRCh37) protein-coding genes with an annotation window of 50 kb. We used the computed gene P-value to calculate the *P_G_* term in GPScore.

#### eQTL colocalization (coloc)

We used a combination of SuSiE ^21^ and coloc ^27^ to assess for evidence of shared association signals/causal variants between our adiponectin GWAS data and *cis*-eQTLs from GTEx ^37^ using the coloc.susie() function ^35^. The coloc + SuSiE approach has been shown to improve the accuracy of colocalization analyses (coloc only) when multiple causal variants exist within a window ^35^. We extracted the posterior probability of the variant being causal for the shared signal (H4) for each tissue most biologically-relevant to adiponectin and used it in the *S_G_* term for the GPScore.

#### eQTL colocalization (LD-based)

As a supplemental approach to coloc+SuSiE, we further examined for colocalizations between our adiponectin-associated variants and eQTLs in GTEx and in RNA-seq data from 434 METSIM participants using an LD-based approach ^26^. For colocalization with the GTEx data (restricted to adiponectin-related tissues described above), we considered the GWAS and eQTL signals to be colocalized when the lead or index GWAS variant and the variant most strongly associated with the expression level of the corresponding transcript (eSNP) exhibited high pairwise LD (LD *r*^2^ > 0.70) in European + East Asian ancestry data within 1KGp328. For colocalization with the METSIM RNA-seq data, we considered the GWAS and eQTL signals to be colocalized when the lead or index GWAS variant and the eSNP exhibited high pairwise LD (LD *r*^2^ > 0.80) calculated using 10,197 METSIM participants. The P-value of association with the eSNP was used to calculate the *P_G_* term in GPScore.

#### Summary-based Mendelian randomization

Summary-based Mendelian randomization (SMR) is another approach that integrates GWAS and eQTL data to identify genes whose expression levels are associated with a complex trait. We applied multi-SNP SMR ^28^ to test for the effect of gene expression (adiponectin-related tissues from GTEx) variation on adiponectin. We included the probes with at least one cis-QTL at *P*_EQTL_ < 5 × 10^−8^, and we performed a HEIDI (Heterogeneity in dependent instruments) test to exclude results that may reflect linkage. In GPScore calculations, we considered the *P_SMR_* values of each probe corresponding to tissue, excluding probes with strong evidence of heterogeneity (*P_HEIDI_*> 0.01). We used the *P_SMR_*to calculate the *P_G_* term in GPScore.

#### SNP-heritability enrichment

To evaluate whether the variant heritability was enriched in the variants within tissue-specific genes (±100 kb) compared to other regions, we applied partitioned LD score regression (LDSC) ^29^ with the ‘-overlap-annot’ option. We constructed the annotation list of tissue-specific genes by selecting genes with the top 10% median TPM (transcripts per million) values in each adiponectin-relevant tissue from the expression data. We calculated the P-values from one-sided Z-score coefficients for tissue-specific genes and included them in the *P_G_* term in GPScore.

#### EMS annotation

The EMS annotation is defined as the predicted probability that a variant has a *cis*-regulatory effect on gene expression, which we calculated by training a random forest model on fine-mapped eQTLs and 6,121 features, such as epigenetic marks and sequence-based neural network predictions ^30^. We extracted the computed EMS for all top variant-gene pairs in adiponectin-relevant tissues in GTEx v8^37^ from the Finucane Lab (https://www.finucanelab.org/data). We then extracted the normalized EMS score for all variants from the adiponectin meta-analyses (*P* < 0.05 and minor allele frequency> 0.50%) and then incorporated it into the *S_G_*term in GPScore.

#### PoPS

A gene-level Polygenic Priority Score (PoPS) is a similarity-based gene prioritization approach that leverages both polygenic and locus-specific genetic signals ^31^. We used PoPS v0.2 to identify potential target genes from gene-level association statistics (derived from MAGMA) by integrating 57,543 gene features from public bulk and single-cell expression datasets, protein-protein interaction networks, and pathway databases. The PoPs score for each gene is then incorporated into the *S_G_* term in GPScore.

#### Downstreamer

We used Downstreamer ^32^ to calculate co-regulation Z-scores per gene using expression data from 31,499 public RNA-seq samples from many different tissues ^38^. Integrating this information into the calculation of the gene priority score will likely prioritize genes that are co-regulated with many important adiponectin-associated genes in the GWAS. We calculated the P-values from one-sided co-regulation Z-scores per gene and included them in calculating the *P_G_* term in GPScore.

#### Epimap

Human epigenome reference, EpiMap (epigenome integration across multiple annotation projects) uses the correlation of enhancer activity with gene expression across cell types to map regulatory SNPs to their target genes. This information is useful in the complex trait investigation and studies of disease locus mechanisms. We downloaded gene-enhancer links by mark activity-by-gene expression correlation for the adipose tissue from the Epimap repository ^11^ and mapped the link scores for the genes overlapping in the 3Mb region around the lead variant. We then incorporated the link scores into the *S_G_* term in GPscore.

#### GeneHancer

GeneHancer calculates a score derived from: gene-enhancer genomic distances and combined scores of tissue co-expression correlation between genes and enhancer RNAs, enhancer-targeted transcription factor genes, eQTLs for variants within enhancers, and capture Hi-C ^18^. The score represents the strength of enhancer association to a particular gene (GeneHancer score), which can be useful in predicting regulatory elements and their target genes. We extracted the GeneHancer scores for enhancer gene pairs for genes physically located within the adiponectin fine-mapping regions and incorporated into the *S_G_* term in GPscore.

#### Combined SNP-to-gene score annotation

The SNP to Gene (cS2G) strategy is a heritability-based framework that combines different SNP-to-gene strategies to link regulatory variants to their target genes. We extracted the cS2G scores, which include 10 main functional SNP-to-gene strategies, as previously described ^33^, from 9,997,231 variants in the 1000 Genomes Project European reference panel and 19,476,620 variants in the UK Biobank. The cS2G score annotation was performed on selected variants (*P* < 0.05 and minor allele frequency > 0.50%) from the meta-analyses. The cS2G score ranges from 0 to 1, where a score>0.5 is considered good evidence; we incorporated this score into the *S_G_* term in GPScore.

### Disease enrichment and Functional association networks

#### Disease enrichment analysis of prioritized genes

To study mechanisms underlying human diseases related to adiponectin, we investigated the human gene-disease associations of our adiponectin-prioritized genes. We examined for enrichment between the prioritized genes from the multi-ancestry adiponectin meta-analysis and a wide range of disease phenotypes relevant in human genomics by gene-disease enrichment analysis (DisGeNET) ^39^, using enrichr ^40^. We considered a DisGeNET term enriched if adjusted *P* < 0.01.

#### Functional association network construction

To investigate potentially functionally-similar genes and their underlying pathways, we constructed a functional association network using GeneMANIA ^41^. Prioritized genes alone do not contain enough information to build networks that mediate the underlying functional relationship. We used an algorithm ^42^ in GeneMANIA that constructs network weights based on the reproducibility of Gene Ontology (GO) biological process co-annotation patterns. We expanded the gene list with functionally similar genes from multiple genomics and proteomics data sources. We primarily used the physical and genetic interaction, predicted protein interaction, and pathway and molecular interaction data available in GeneMANIA.

## Results

### Multi-ancestry meta-analysis reveals 15 known and 7 novel risk loci for adiponectin

We conducted a multi-ancestry, genome-wide association meta-analysis for adiponectin using summary statistics from the METSIM cohort ^15^ and the ADIPOGen ^4^ and AGEN consortia ^5^ (*N* ≤ 46, 434; Table S1). We also performed a genome-wide association meta-analysis for adiponectin in data from up to 38,609 individuals of European-ancestry (METSIM cohort and ADIPOGen consortium only) ^4, 15^. Each individual cohort performed single-variant association analyses for adiponectin, adjusted for age, sex, BMI, and study-specific covariates as appropriate (e.g., principal components, study site). We meta-analyzed summary statistics using fixed-effects analyses implemented in METASOFT^16^. We corrected for population substructure in the multi-ancestry meta-analysis using the computed inflation factors for both the mean effect (λ_MeanEffect_) and heterogeneity portions (λ_heterogeneity_) ^16^. Because RE2^16^ and MR-MEGA ^17^ are not sufficiently powered in a meta-analysis of only three input files ^43^, we consider the fixed-effects results as our primary results; however, we present results from RE2 and MR-MEGA for comparison.

#### Multi-ancestry meta-analysis association results

We identified 22 loci associated with adiponectin (*P*<5×10^−8^), including seven loci that have not previously been associated with adiponectin (Table 1, Figure 1, Figure S1). Previously unreported loci are located at or near *CSF1*, *RGS17*, *ADRB1*, *PDE3B*, *RBMS2*, *HCAR1*, and *PHF23*. Effect direction for all lead variants showed concordance between all three studies, and the effect sizes for each of the lead variants were also consistent between the three datasets (Spearman r≥ 0.88; Figure S2). Further, MR-MEGA results for all 22 loci have a log10BF > 30, further supporting the results from the fixed-effect models. We also confirmed (*P*<0.05) 83 out of 167 (49.7%) previously reported variants to be associated with adiponectin in our meta-analysis (Table S2).

**Figure 1.**
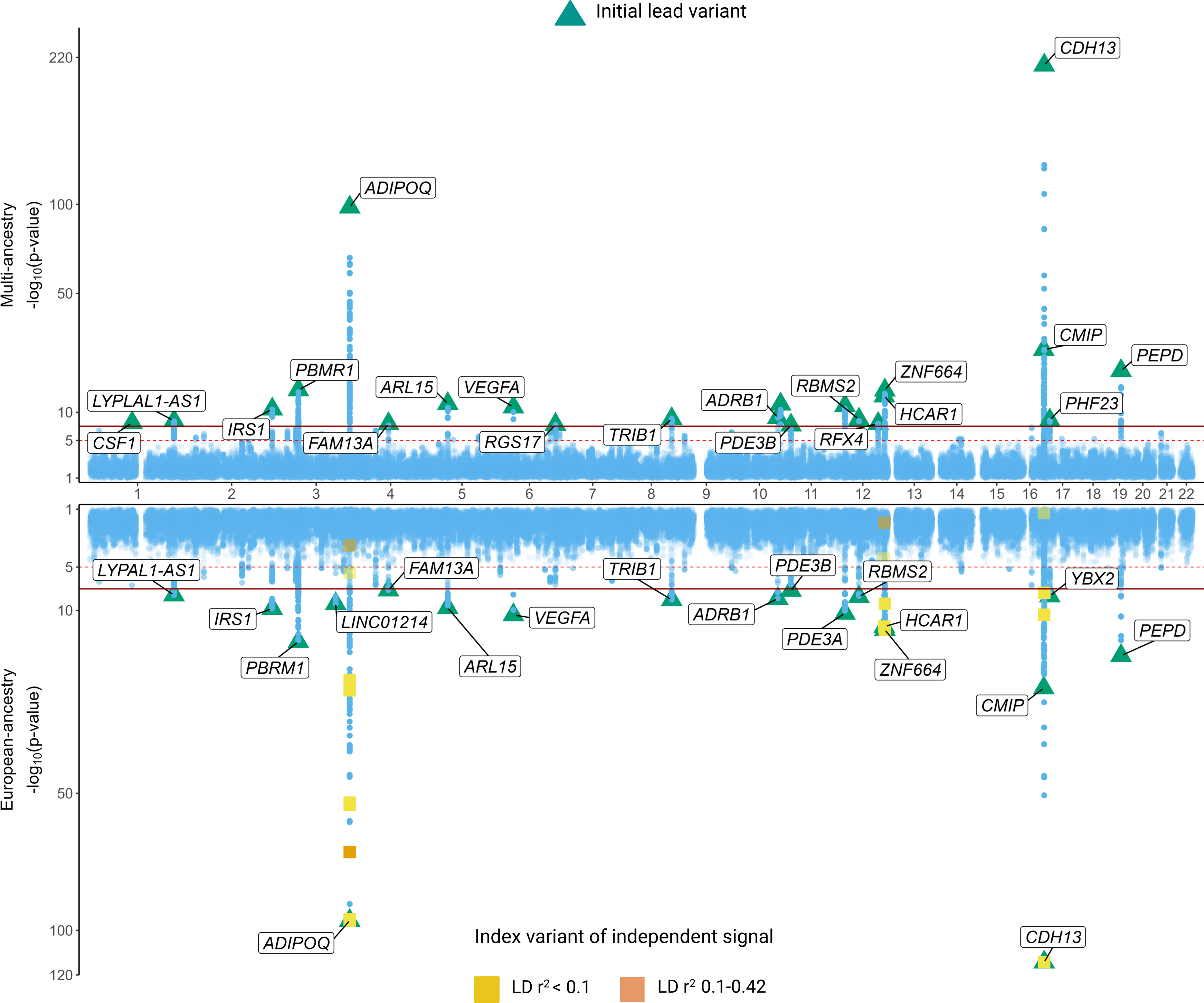
Miami plot of the multi-ancestry (top) and European-ancestry (bottom) genome-wide meta-analysis results for adiponectin. The y-axis represents −log10(P) value from the fixed-effects analyses. Each blue dot represents a variant tested in the meta-analysis. The transcript closest to the lead variant (triangles) at each locus is listed in rectangle boxes. Index variants from the additional distinct association signals identified in the European-ancestry meta-analyses are denoted by squares and colored based on the linkage disequilibrium with the lead variant at that locus (orange: 0.10<r^2^<0.42; yellow: r^2^<0.10).

**Table 1.**
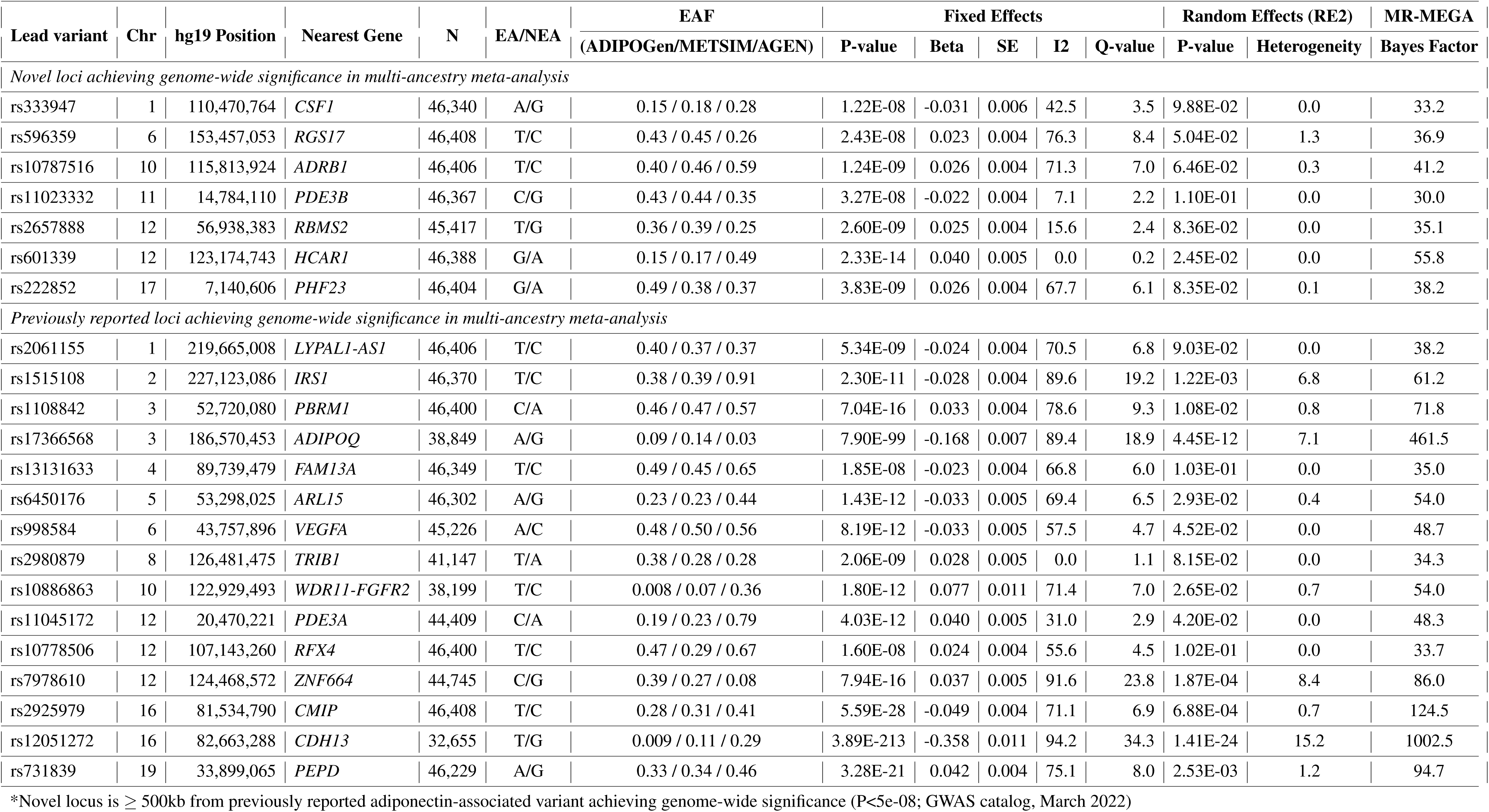
Multi-ancestry meta-analysis of adiponectin associations. . Lead variant at each locus attaining genome-wide significance (*P <* 5 *×* 10*^-^*^8^) in the multi-ancestry meta-analysis (ADIPOGen, AGEN, METSIM).

Lead variants at four loci exhibited evidence of heterogeneity in effect sizes (*I*^2^ > 80%; rs1515108 (*IRS1*), rs17366568 (*ADIPOQ*), rs7978610 (*ZNF664*), rs12051272 (*CDH13*); Table 1). All four loci are well-established associations with adiponectin levels ^4, 5, 7^. Among the four heterogeneous loci, two of the loci were still considered statistically significant in RE2 models accounting for the high heterogeneity (rs17366568, rs12051272). Large differences in effect allele frequencies between European- and East-Asian ancestry populations is likely driving the observed heterogeneity (Table 1, Figure S2).

#### European meta-analysis

In a meta-analysis of European-ancestry individuals from METSIM and the ADIPOGen consortium, we identified 19 loci associated with adiponectin (Table 2, Figures 1, S2), 18 of which were also identified in the multi-ancestry results. One additional previously unreported locus, located near *LINC01214* on chromosome 1 (rs7617025) was identified only in the European-ancestry analysis. All the lead variants showed effect direction concordance between METSIM and ADIPOGen.

**Table 2.**
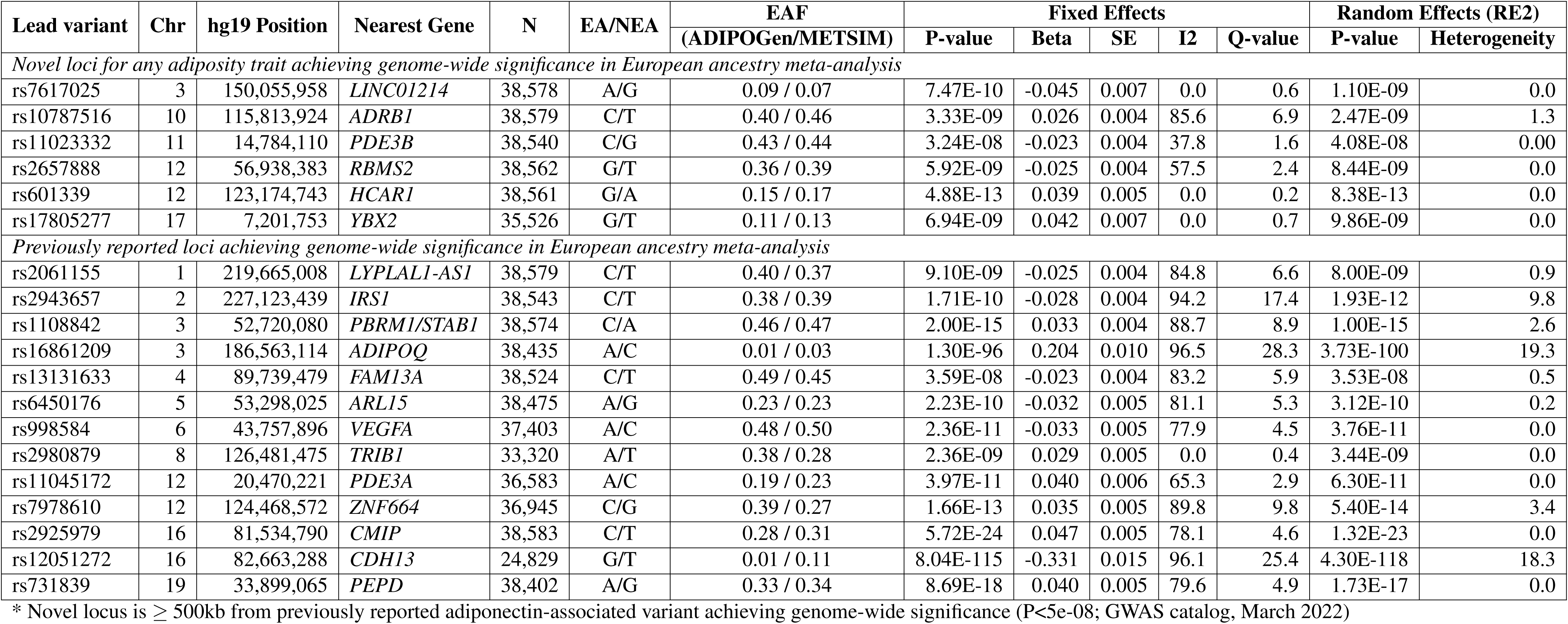
Meta-analysis of adiponectin associations in European-ancestry population. . List of all lead variants achieving genome-wide significance (*P <* 5 *×* 10*^-^*^8^) in the meta-analysis of adiponectin associations in the European population (ADIPOGen and METSIM).

#### Distinct association signals

At all 19 loci identified in the European-ancestry, we sought to identify additional association signals located within +/- 1 Mb of the lead variant using GCTA-COJO. We detected (*P* < 5 × 10^−8^) 14 additional signals at four loci (Table S3; Figures 1, S3), including eight signals near *ADIPOQ* (LD *r*^2^ = 0.001-0.846), one near *HCAR1* (LD *r*^2^ = 0.183), one near *ZNF664* (LD *r*^2^ = 0.008), and four near *CMIP*/*CDH13* (LD *r*^2^ = 0-0.046) (Table S4).

#### Pleiotropic associations with complex traits

We evaluated the lead variants at each of the 22 loci reported in the multi-ancestry meta-analysis for association with other complex traits and diseases. For these traits, we examined existing GWAS and genome-wide meta-analysis results in the Common Metabolic Diseases Knowledge Portal (www.cmdkp.org). We found 89 unique lead variant-phenotype combinations that reached Bonferroni significance (Table S5). Lead adiponectin-associated variants are most strongly associated with the lipids, hepatic, hematological, glycemic, and anthropometric phenotype groups (Figure S4, Table S5).

### Putative causal variants implicate shared biological mechanisms

#### Fine-mapping to identify potential causal variants

To identify potential causal variants underlying adiponectin association signals in both the multi- and European-ancestry analyses, we first constructed 3 Mb windows (+/- 1.5 Mb) around the lead variant (Figure S5). The variants with a PIP> 0.90 in either FINEMAP ^20^ or SuSiE ^21^ and LD *r*^2^ > 0.8 with the lead GWAS variant were identified as candidate causal variants. In fine-mapping the multi-ancestry adiponectin loci, we nominated 46 putative causal variants at 17 loci (Table S6; Figure S6). Among the 46 putative causal variants, 10 (rs2061155, rs1515108, rs1108842, rs998584, rs596359, rs10787516, rs10886863, rs7978610, rs12051272, rs731839) were also the lead variants from the meta-analysis. In the European-ancestry analyses, fine-mapping analyses nominated putative causal variants at seven loci, including two variants near *CDH13* (Table S7; Figure S7). Among the seven putative causal variants, six (rs16861209, rs998584, rs11045172, rs2925979, rs12051272, rs731839) were the lead variants from the meta-analysis.

#### Regulatory effects of nominated causal variants

To investigate the regulatory effects among our nominated causal variants, we used the RegulomeDB ^23^ to obtain RegulomeDB scores, which rank the presence of regulatory motifs for variants on a scale from 1a (most evidence) to 7 (no data). In multi-ancestry fine-mapping results, 17 of the 46 putative causal variants were scored as “regulatory” (score < 5, with probability prob≥ 0.50; Table S6). rs683039, located at 12kb 5’ of *RGS17*, had the lowest RegulomeDB score (score=1f; prob=0.93), demonstrating the largest evidence for being in a regulatory region (Figure S1). In European-ancestry fine-mapping results, six of the eight putative causal variants were scored as “regulatory” (score < 5, with prob p≥ 0.50; Table S7). rs2925979, located near *CMIP*, had the lowest RegulomeDB score (score=2b; prob=0.75), demonstrating the largest evidence of being in a regulatory region.

#### Causal variants enriched for cardiometabolic traits

We also examined our nominated causal variants for shared genetic architecture with fine-mapping results of 2,629 unique traits from over 3,052 GWAS summary statistics obtained from the CAUSALdb database ^24^, which includes UK Biobank and other cohorts. When a candidate adiponectin-causal variant was reported in more than one prior study, we meta-analyzed the p-values. In total, we found 306 unique lead variant-trait combinations that are Bonferroni significant (Table S8, Figure S8). We observed that most of our nominated causal variants show prior strong associations for liver-related diseases, type 2 diabetes mellitus, hormones, body size, and body composition measurements (Figure S8).

### Gene prioritization highlights genes underlying adiponectin association signals

#### GPScore to nominate locus-specific target genes

Variants underlying genetic association signals may not necessarily regulate the closest gene ^44–48^. To assess and prioritize target genes underlying adiponectin-associated signals, we developed a combinatorial likelihood scoring formalism (“GPScore”) based upon measures derived from 11 gene prioritization strategies (Tables S9-S30), along with the physical distance to the transcription start site(s) (TSS) (Figure 2). We integrated multiple lines of evidence into the score by using four terms: *P_G_* (combined p-value), *S_G_* (combined scaled scores), *C_S_* (proportion of support), and *TSS_d_* (distance from TSS of the gene to the lead GWAS variant). The most likely target gene(s) were selected based on the highest locus-specific GPScore within the 3Mb window. For the four regions with overlapping windows (12q24.31 near *HCAR1* and *ZNF664* and 16q23 near *CMIP* and *CDH13*), the flanking size of 1.5 Mb is reduced to 1 Mb to limit the window overlap.

**Figure 2.**
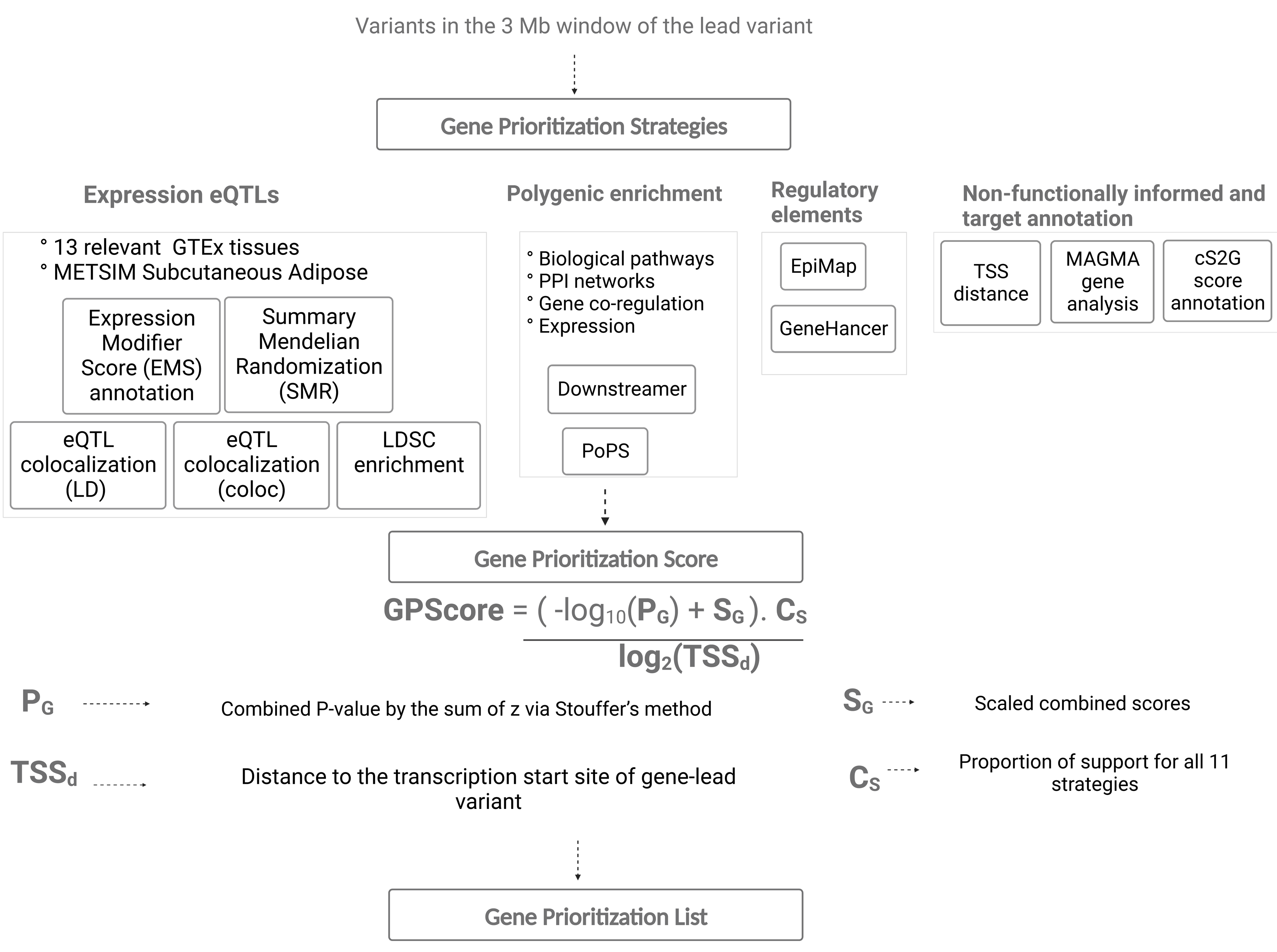
Schematic overview of gene prioritization score construction for finding target genes of adiponectin. Flow diagram illustrating various gene prioritization strategies using multiple data sources: expression QTLs, pathways and networks, regulatory elements, and non-functionally informed methods. The gene prioritization score incorporates all this evidence using four terms: PG, SG, CS, and TSSd (distance to the transcription start site). We prioritize locus-specific target genes using the gene prioritization score, with higher values indicating stronger support across approaches.

In multi-ancestry analysis, we prioritized a total of 30 genes with high relative locus-specific GPScore values across 22 locus windows (Table 3, S31; Figure 3). Out of 22 regions, 15 had one prioritized gene, six had two prioritized genes (*PBRM1* and *GNL3* at 3p21.1, *ADIPOQ* and *RFC4* at 3q27.3, *NAP1L5* and *FAM13A* at 4q22.1, *FDGFR2* and *WDR11* at 10q26.12, *CTDNEP1* and *ELP5* at chr17p13.1, and *PEPD*, and *CEBPG* at 17p13.1), and one region had three prioritized genes (*CCDC92*, *DNAH10OS*, and *ZNF664* at 12q24.31) (Table 3, Figure 3). GPScore prioritized well-known adiponectin target genes concerning various cardiovascular/cardiometabolic diseases including *IRS1*, *ADIPOQ*, *PDE3A*, *CCDC92*, *CDH13* ^49–53^. GPScore also prioritized target genes for previously unreported loci that have been shown to play key roles in adipose-related metabolic functions (e.g., *CSF1*, *RGS17*, *ADRB1*, and *CYP2R1*) ^54–57^.

**Figure 3.**
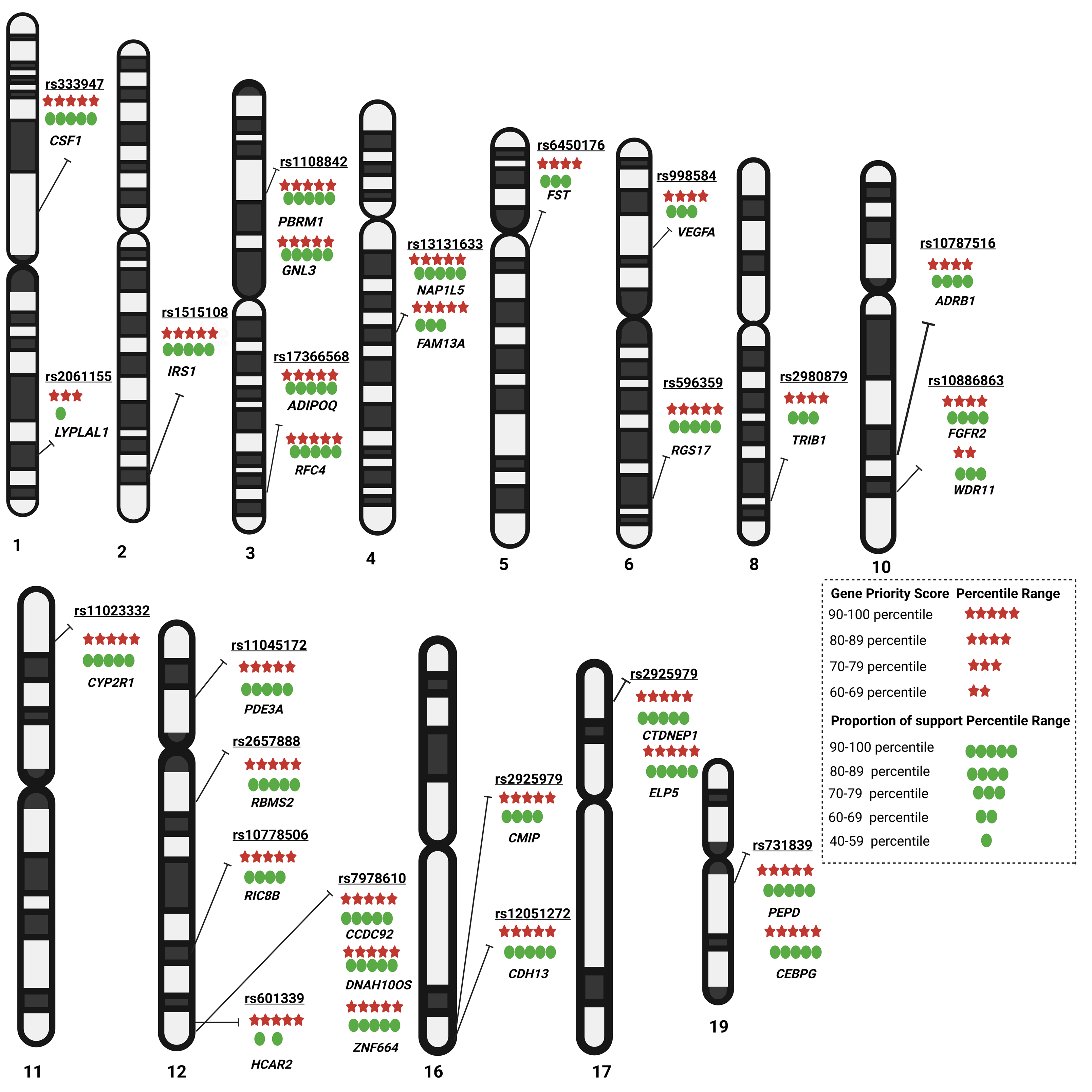
Adiponectin locus-specific prioritized genes and strength. A chromosome ideogram depicts the location and strength of GPScore values, CS (proportion of support) of each prioritized gene identified in an adiponectin multi-ancestry meta-analysis. Red stars highlight the strength of the GPScore measured in percentile, whereas green circles highlight the proportion of support from the 11 prioritization approaches. Some locations have multiple prioritized genes with relatively similar scores.

**Table 3.**
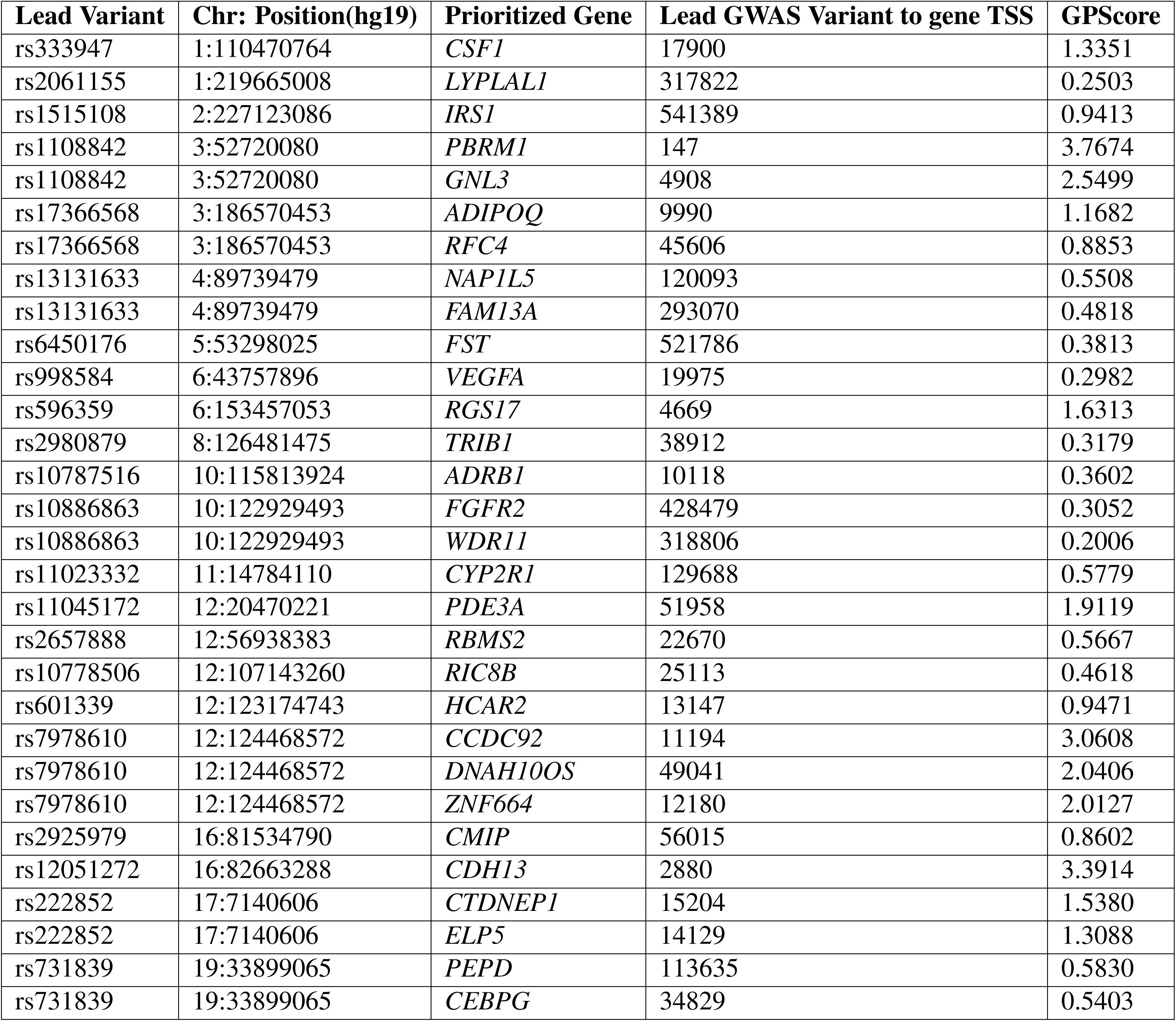
Adiponectin multi-ancestry meta-analysis prioritized target gene list. Prioritization of 30 likely target/effector genes by using locus-specific gene priority score constructed from eleven gene prioritization strategies along with TSS distance.

Gene prioritization results using variants from the European-ancestry analysis yielded similar results to the multi-ancestry prioritization (Table S32). At the 18 loci shared between European- and multi-ancestry analyses, 13 loci yielded the same set of prioritized genes. At 4 of the shared loci, the European-ancestry GPScore prioritized additional target genes (Table S32). At the chr17p13.1 locus shared between the European- and multi-ancestry results, GPScore prioritized a different target gene in the European-ancestry analysis (*ACAP1*) than in the multi-ancestry analysis (*CTDNEP1* and *ELP5*). Differences in GPScore prioritization between the two sets of analyses is likely explained by differences in effect sizes and allele frequencies used as input to the gene prioritization strategies.

To see which of the GPScore terms received the most weight in the score calculation, we quantified each term’s contribution to the GPScore (Figures 4a, S9) by fitting a multiple regression model and averaging over orderings proposed by Lindeman et al. and implemented in the *realimpo* package^58^. Across the 22 multi-ancestry regions tested, the average contributions (on a scale of 0-1) were: *S_G_*=0.133, *C_S_*=0.199, *TSS_d_*=0.255, and *P_G_*=0.411. The 2q36.3 window where *IRS1* is prioritized has the lowest *TSS_d_*contribution since the transcription start site for *IRS1* is 541,389bp away from the lead adiponectin GWAS variant but has the strongest support for being the effector gene (Table S31). The 3p21.1 window has the highest *TSS_d_* contribution since both the prioritized genes *PBMR1* and *GNL3* are close to the lead variant (Figure 4d). The variants in 5q11.2,6p21.1 and 8q24.13 windows have the lowest contribution of *C_S_*(proportion of support) due to a lack of support from many of the gene prioritization strategies (Figure 4a). The *P_G_* (combined p-value) has an average contribution of 0.411 across all the windows. Regarding the gene prioritization strategies, all prioritized genes have MAGMA P-value < 5×10^−8^, relatively higher POPS scores, and CS2G annotations while supporting the majority of other strategies (Figure S9). Because several prioritization strategies rely on a reference panel for LD calculation, we repeated GPScore using only the METSIM cohort summary statistics; the top prioritized genes for the common windows remain the same (Table S33).

**Figure 4.**
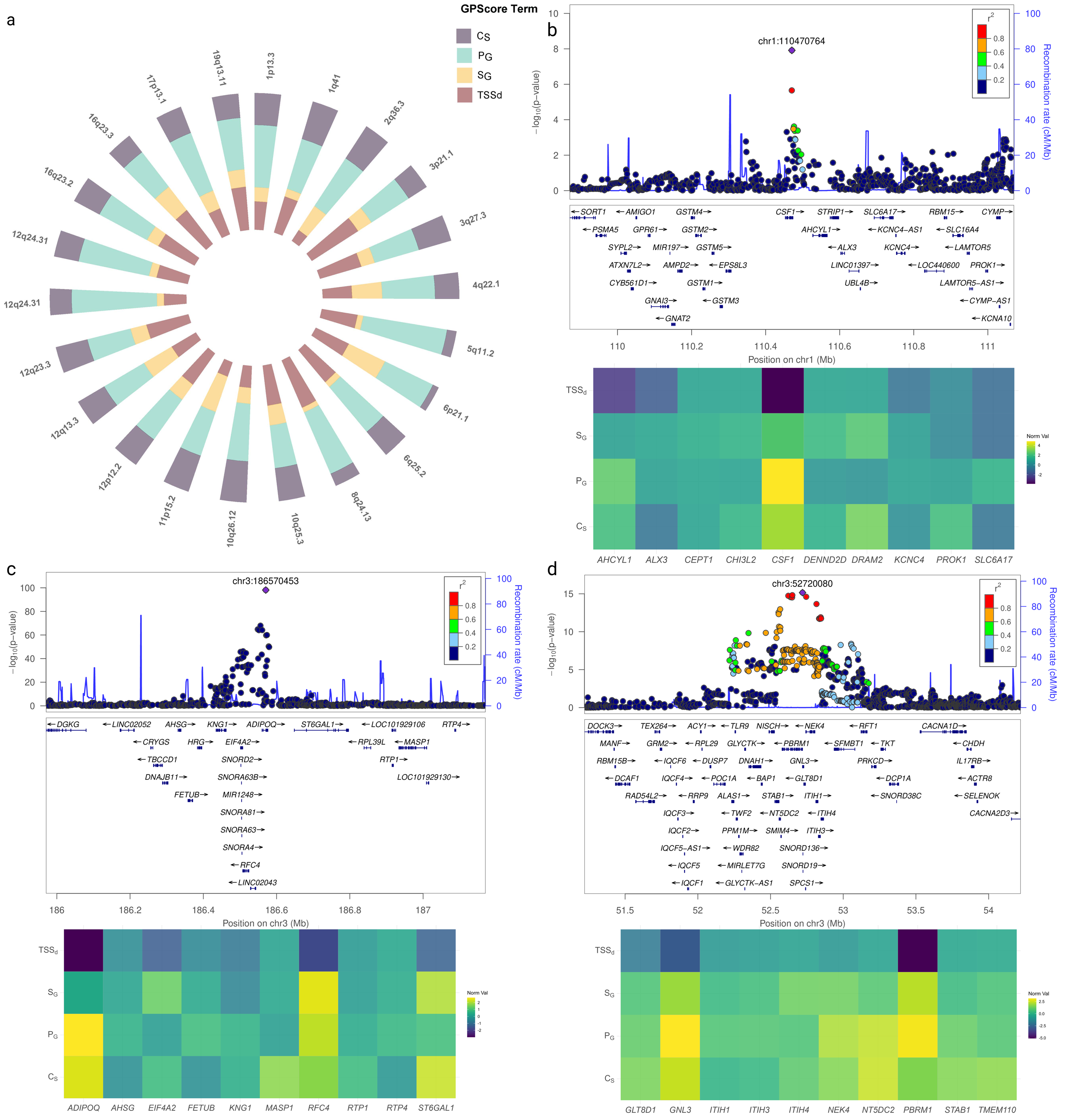
The relative contribution of terms from the Gene Priority Score and illustrative biological examples. a. Relative importance of four terms: PG, SG, CS, and T SSd for the gene priority score construction across 22 associated regions in multi-ancestry analysis. b-d. Top: Locus zoom plots of multi-ancestry summary statistics, colored by LD with the lead variant. The color of the heatmap represents enrichment for the GPScore term. The top prioritized genes at each exemplary locus are CSF1 (b), ADIPOQ and RFC4 (c), GNL3 and PBRM1 (d).

### Pinpointing biological functionality through disease enrichment and functional interaction networks

#### Disease enrichment analysis

We evaluated the association of multi-ancestry prioritized genes with disease phenotypes using gene-disease enrichment analysis ^39, 40^. We found a strong enrichment (adjusted *P* < 0.01) of DisGeNET terms related to cardiometabolic traits and diseases, including waist-hip ratio, arteriosclerosis obliterans, non-alcoholic fatty liver disease, body mass index, coronary artery disease, hypoadiponectinemia, and high-density lipoprotein measurement (Figure 5a, Tables S34-35). Other highly enriched terms are related to cancer outcomes, including thymoma, endometrial neoplasms, endometrial adenocarcinoma, giant cell tumors, noninfiltrating intraductal carcinoma, and non-hereditary clear cell renal cell carcinoma. These findings support recent epidemiological evidence that links adiponectin to cancer ^59, 60^.

**Figure 5.**
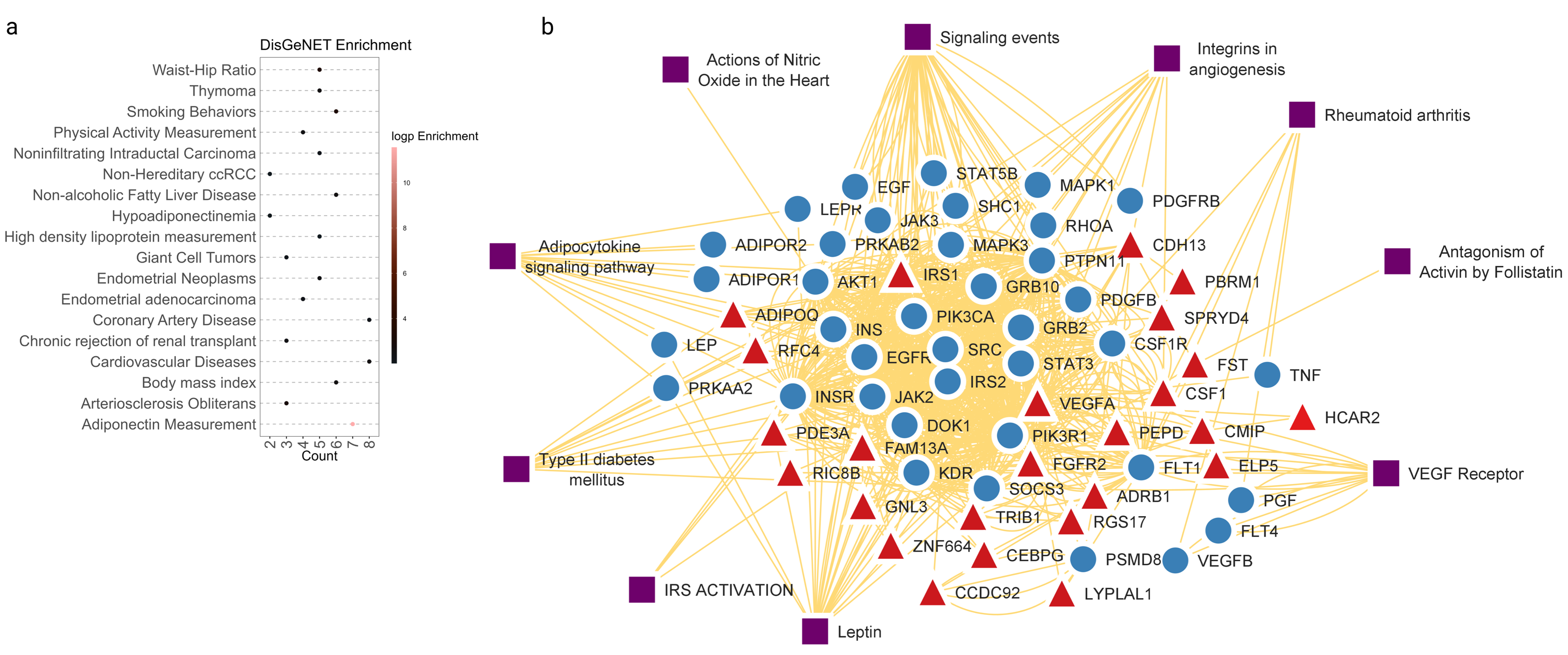
Disease enrichment and functional interaction network of adiponectin multi-ancestry prioritized genes. a. Select results from the disease enrichment analysis (DisGeNET, enrichr) using the multi-ancestry prioritized genes list. DisGeNET terms related to cardiovascular/cardiometabolic traits show strong enrichment (adjusted p < 0.01). b. An interactive functional association network, illustrating the relationships among the connected genes (blue circles), prioritized genes (red triangles), and underlying associated pathways constructed using GeneMANIA. A total of 10 pathways were represented in the network (purple squares). A complete network is shown in Figure S10.

#### Functional association network

For the multi-ancestry meta-analysis results, we constructed an interactive functional association network, illustrating the relationships among the connected genes, prioritized genes, and associated pathways (Figures 5, S10; Tables S36-39). In Figure 5, we display the network results where we use the default of adding 20 “related” genes and pathways into the network. With the default network size, 76.6% of our prioritized genes (23/30) were incorporated into functional association networks, with most having multiple connected genes (Figure 5b). A total of 10 pathways were represented in the network, with many genes heavily involved in the adipocytokine signaling pathway, integrins in angiogenesis, leptin pathway, and type 2 diabetes.

## Discussion

Adiponectin is an insulin-sensitizing and anti-inflammatory hormone secreted by adipocytes whose signaling is involved in several metabolic processes crucial to type 2 diabetes, neurodegeneration, and cardiovascular disease. In this study, we conducted the largest multi-ancestry adiponectin GWAS meta-analysis to date, in up to 46,434 individuals from the METSIM cohort and the ADIPOGen and AGEN consortiums. We identified 22 loci associated with adiponectin (*P* < 5 × 10^−8^), including 15 known and seven novel loci. European-ancestry results revealed 14 additional secondary signals at the *ADIPOQ*, *CDH13*, *HCAR1*, and *ZNF664* loci. Most of the lead variants showed associations with other complex traits/diseases, suggesting pleiotropy. We narrowed the list of putative causal variants to 46 (PP>0.9), 17 of which demonstrated moderate to strong regulatory evidence. Our nominated causal variants also showed potential pleiotropy for other complex cardiometabolic traits, such as liver-related diseases, type 2 diabetes, hormones, body size, and body compositional measurements. Variants underlying genetic association signals may not necessarily regulate the closest gene ^44–48^ and may impact protein levels thousands or even millions of base pairs away ^61^. Based on prior estimates ^62^, the 3 Mb fine-mapping mapping region (+/- 1.5 Mb around the index variant) should roughly capture more than 99% of such regulatory activity. Multiple methods currently exist to prioritize target, or effector, genes underlying GWAS signals, primarily from gene expression quantitative trait loci (eQTL) data ^10^, regulatory genomic activity ^11^, and protein-protein interaction networks and pathway databases ^12^. Two common challenges of existing gene prioritization approaches are 1) inadequate customizability to represent the data which is disease/tissue relevant, and 2) conflicting findings from various approaches due to differences in assumptions, methodology, and representation models, resulting in the selection of methods/results in an ad hoc or post hoc manner. To address these limitations, we developed combinatorial likelihood scoring formalism, GPScore, based upon measures derived from 11 gene prioritization strategies, along with the physical distance to the transcription start site(s) (TSS). GPScore integrates multiple lines of evidence into the unbiased score that can be customizable to include weighting factors to prioritize signals from a particular tissue and can be broadly applicable to other complex traits that do not have much training data. By utilizing GPScore, we prioritize the 30 most probable target/effector genes for adiponectin in the multi-ancestry analysis. The prioritized gene list includes several well-known causal genes related to serum adiponectin levels. This list includes *ADIPOQ*, which is responsible for encoding the adiponectin protein, and *CDH13*, which encodes an adiponectin receptor protein ^51^. Insulin receptor substrate-1 (*IRS1*) is a substrate of the insulin receptor tyrosine kinase, and it is central to inducing insulin actions, including binding and activating PI3K and increasing glucose transport ^63^. Acute knockdown in adipocytes was shown to result in increased Adipoq mRNA expression, but reduced adiponectin secretion ^64^. The treatment of myoblast cell line (C2C12) with adiponectin was also shown to enhance the ability of insulin to stimulate *IRS-1* tyrosine phosphorylation and Akt phosphorylation ^50^. *CCDC92* has been shown to be important in insulin resistance and influencing adipocyte differentiation ^65^, and knockdown of *CCDC92* in mice was shown to reduce obesity and insulin resistance ^66^. We also used GPScore to prioritize target genes among our novel loci from our multi-ancestry meta-analysis, including several for which the biological mechanisms underlying the association with adiponectin may be unclear. The *CSF-1* gene, which was prioritized at 1p13.3, has shown to have therapeutic potential in tissue repair ^67^ due to its involvement in macrophage homeostasis and inflammation via the *CSF-1* receptor ^68^. There is also some supportive evidence that *CSF-1/CSF-1R* signaling is essential in the pathology of and diabetes ^69^. Due to its essential role in G protein signaling, *RGS17* is considered a potential therapeutic target for lung and prostate cancers ^70^; however, individuals with type 2 diabetes have been shown to have higher levels of *RGS17* auto antibodies than individuals without type 2 diabetes, and expression levels of *RGS17* are significantly upregulated in adipose tissue during periods of weight loss ^71^. *ADRB1*, which encodes the adrenergic beta-1 receptor, was prioritized at 10q25.3. While ADRB1 is most known for the functional role it plays in cardiomyocyte function ^72, 73^, visceral adiponectin release was shown to be triggered by cAMP via signaling pathways involving adrenergic beta receptors in mice. The CYP2R1 protein, a receptor for vitamin D 25-hydroxylase in the liver ^74^, plays a crucial role in non-alcoholic fatty liver disease ^75^. Epidemiological studies have also demonstrated associations between serum vitamin D and adiponectin levels. While there is less existing evidence to support the mechanism(s) between some of the prioritized genes and adiponectin levels (e.g., *TRIB1*, *RBMS2*), a further experimental examination into the prioritized genes is warranted.

Our functional profiling of prioritized genes with human disease phenotypes revealed the enrichment of terms related to cardiovascular diseases, coronary artery disease, and other cardiometabolic traits. Interestingly, many genes also showed enrichment related to cancer etiopathogenesis, and recent evidence has shown adiponectin’s antiangiogenic and tumor growth-limiting properties of adiponectin ^59^. Our functional association network analysis also helped illustrate the complex interactions of prioritized genes, their functionally connected genes, and their underlying pathways. We observed the key underlying pathways centered around insulin signaling (through *IRS1*, *IRS2*, *INS*, *INSR*) and adiponectin signaling (thorough *ADIPOR1*, *ADIPOR2*) suggesting potential crosstalk and an essential role in regulating energy balance in the body, inflammation, coagulation, fibrinolysis, insulin resistance, and diabetes.

There are several caveats to our current GPScore gene prioritization approach that should be considered. First, while we were able to utilize our approach in both single- and multi-ancestry summary data, our multi-ancestry analysis is still primarily European (83.14%) and includes only one other ancestry population (East-Asian ancestry); thus, more diverse studies are needed to determine if our approach is less robust when including data with additional heterogeneity. Because our data was largely from European-ancestry populations, we used data from 10,197 Finnish METSIM participants as the reference for LD calculations for our fine-mapping and gene prioritization strategies. We acknowledge, however, that this is likely not the most appropriate panel for LD calculation in our multi-ancestry population. We plan to expand our work by utilizing a larger, more diverse ancestry LD reference panel. However, the flexibility of our approach allows researchers to determine the most appropriate LD panel for their study population. The GPScore prioritization procedure depends on the quality and availability of disease/cell-relevant data, the accuracy of the strategy used, and the annotation quality of variants and genes. As additional data becomes available, it could easily be incorporated into our GPScore approach. It is also possible that transforming variant/position-level scores into gene-level scores may result in a loss of information; however, this information is still retained in the initial prioritization approaches and could still be considered. Lastly, while beneficial to rank genes in a locus-specific manner, the strength of GPScore does not implicate causality for a complex trait or disease, nor is it comparable across locus windows.

In conclusion, we discovered 15 known and seven previously unreported genomic loci associated with adiponectin through multi-ancestry genome-wide meta-analysis. The 46 putative causal variants identified through fine mapping showed potential pleiotropy for cardiovascular/metabolic traits. In addition, we provide a novel and customizable gene prioritization method, GPScore, that integrates multiple lines of evidence, thus increasing the likelihood of identifying the true target gene underlying a GWAS association signal. Our prioritized genes and underlying pathways yield new insights into the genetic architecture of adiponectin. Future research should include phenotyping of adiponectin in large-scale, diverse biobanks and cohorts, thus increasing sample size and ancestral diversity for robust prediction of target genes.

## Data Availability

We will make the final summery stat data available upon publication. IN the interim, all of the summary data of the ADIPOGen consortium, AGEN consortium, and METSIM study are publicly available and may be accessed through their respective websites. The code to reproduce the analyses and figures of this paper is available as Jupyter Notebooks at https://github.com/vsarsani/GPScore.

## Acknowledgements

We thank the individuals in the METSIM cohort for their continued study participation.

## Data and code availability

The summary data of the ADIPOGen consortium, AGEN consortium, and METSIM study may be accessed through their respective websites. The code to reproduce the analyses and figures of this paper is available as Jupyter Notebooks at https://github.com/vsarsani/GPScore.

