## Supplemental Figures for "A multi-ancestry genome-wide meta-analysis, fine-mapping, and gene prioritization approach to characterize the genetic architecture of adiponectin"

### Supplementary Materials

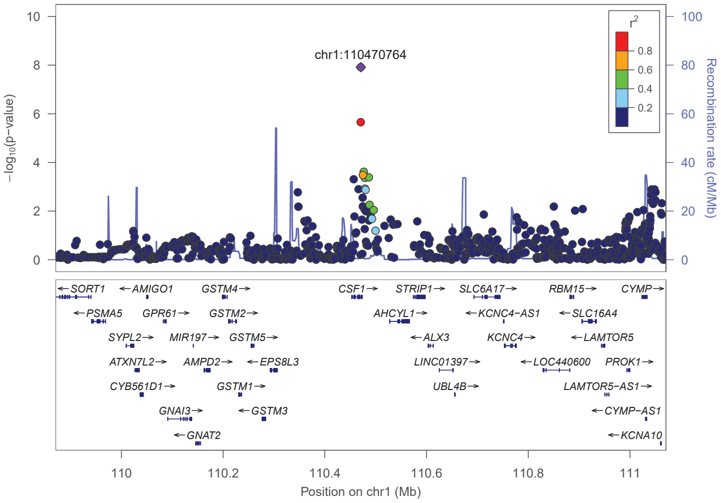


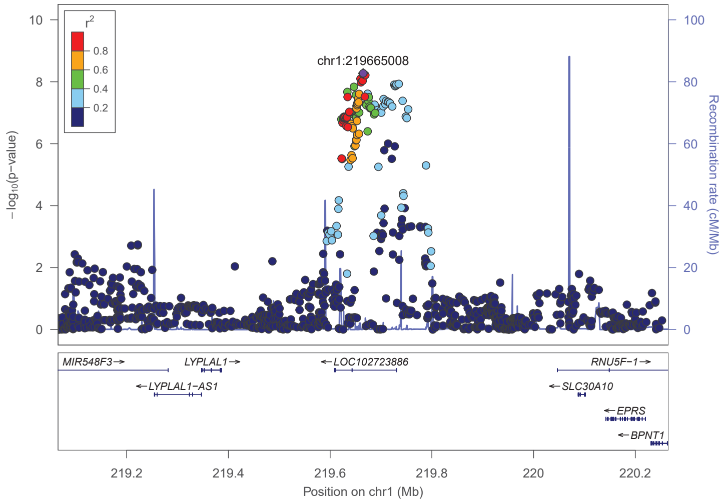


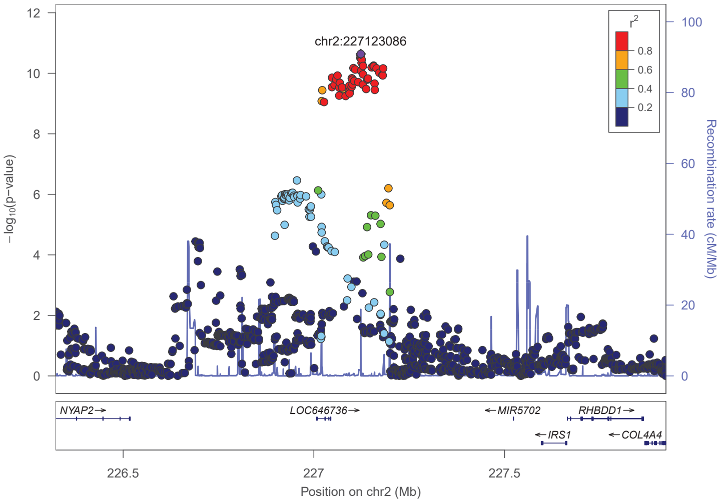


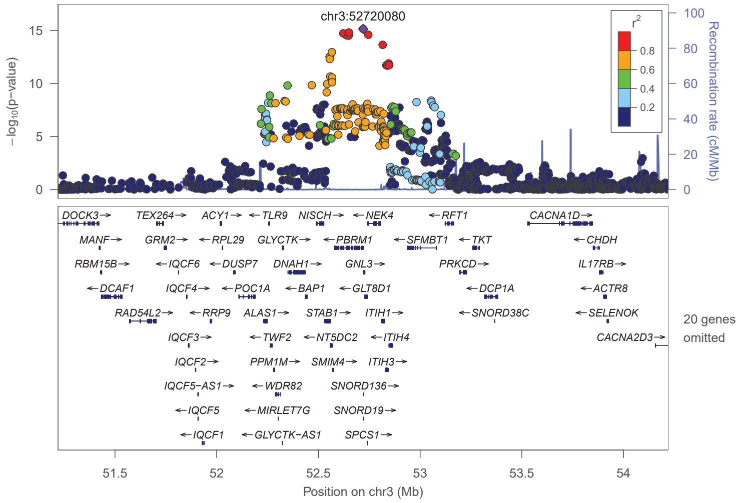


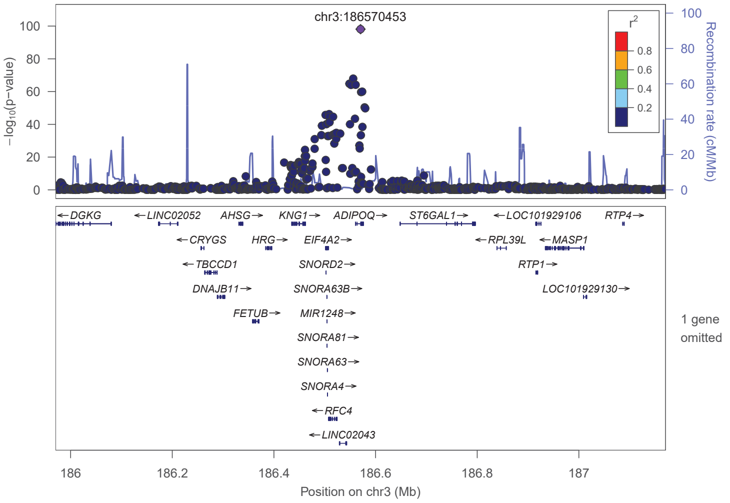


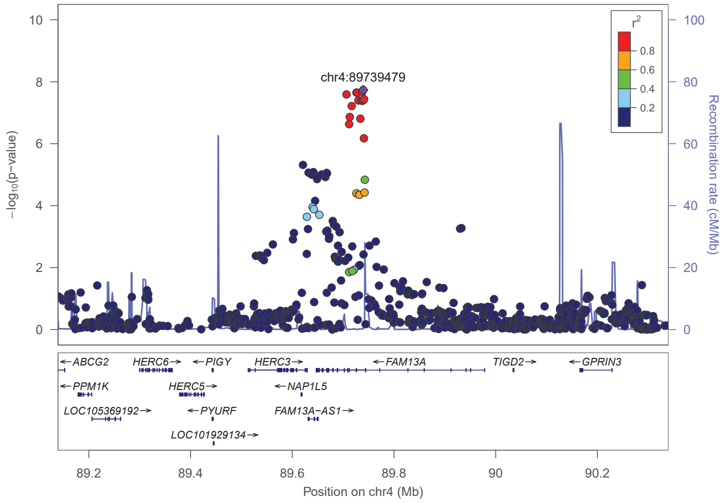

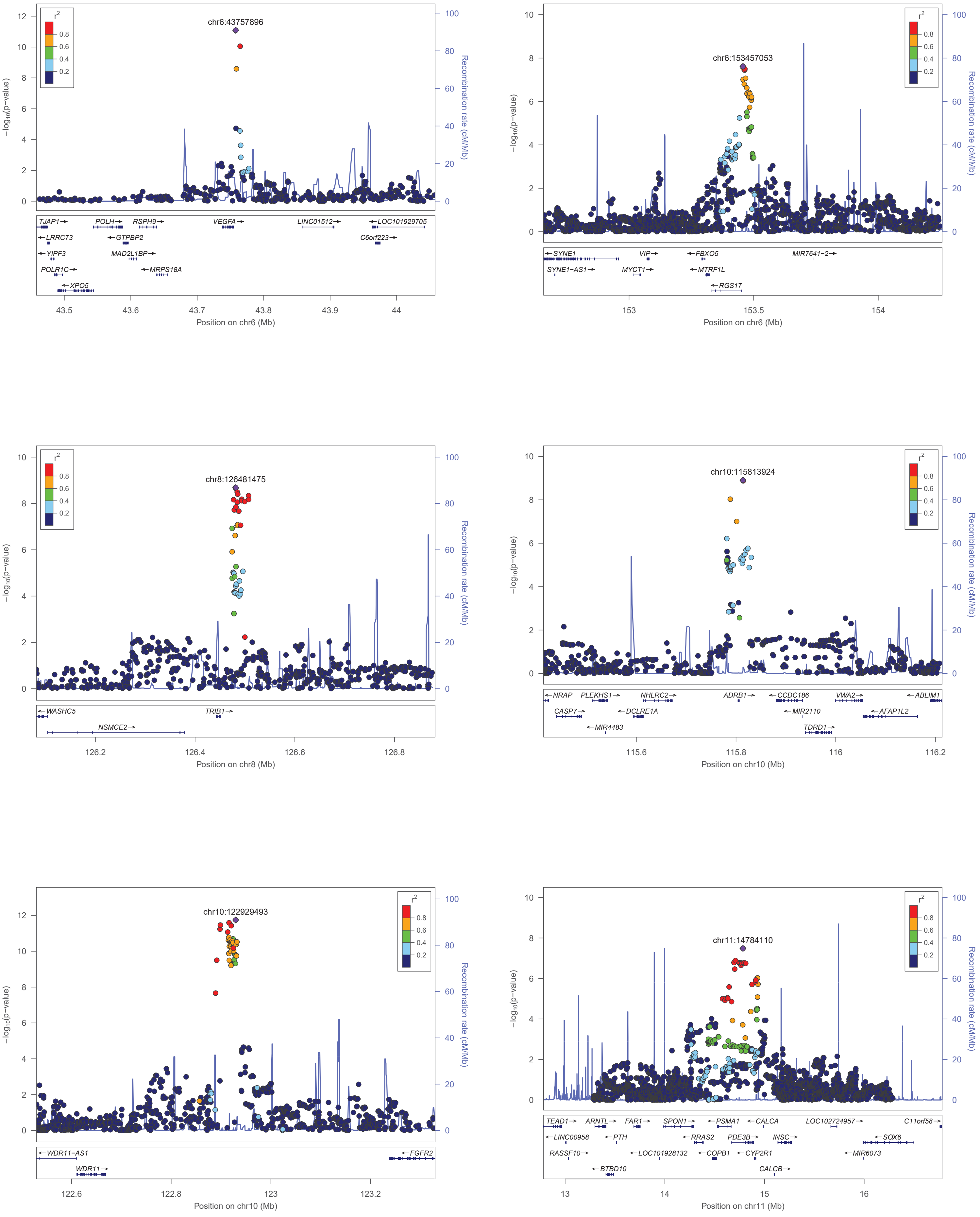

chr6:43757896
−log10(p−value)
Recombination rate (cM/Mb)
r2
TJAP1 LRRC73 YIPF3 POLR1C XPO5 POLH GTPBP2 MAD2L1BP RSPH9 MRPS18A VEGFA LINC01512 LOC101929705 C6orf223
Position on chr6 (Mb)
chr6:153457053
SYNE1 SYNE1−AS1 VIP MYCT1 FBXO5 MTRF1L RGS17 MIR7641−2
Position on chr6 (Mb)
chr8:126481475
WASHC5 NSMCE2 TRIB1
Position on chr8 (Mb)
chr10:115813924
NRAP CASP7 PLEKHS1 MIR4483 NHLRC2 DCLRE1A ADRB1 CCDC186 MIR2110 TDRD1 VWA2 AFAP1L2 ABLIM1
Position on chr10 (Mb)
chr10:122929493
WDR11−AS1 WDR11 FGFR2
Position on chr10 (Mb)
chr11:14784110
TEAD1 LINC00958 RASSF10 ARNTL BTBD10 PTH FAR1 LOC101928132 SPON1 RRAS2 COPB1 PSMA1 PDE3B CYP2R1 CALCA INSC CALCB LOC102724957 SOX6 MIR6073 C11orf58
Position on chr11 (Mb)

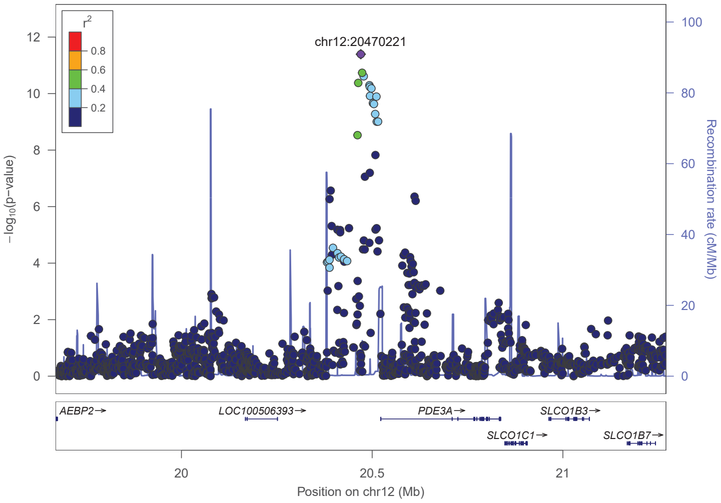
chr12:20470221
r2
0.8
0.6
0.4
0.2
−log10(p−value)
Recombination rate (cM/Mb)
AEBP2→
LOC100506393→
PDE3A→
SLCO1B3→
SLCO1C1→
SLCO1B7→
20
20.5
21
Position on chr12 (Mb)

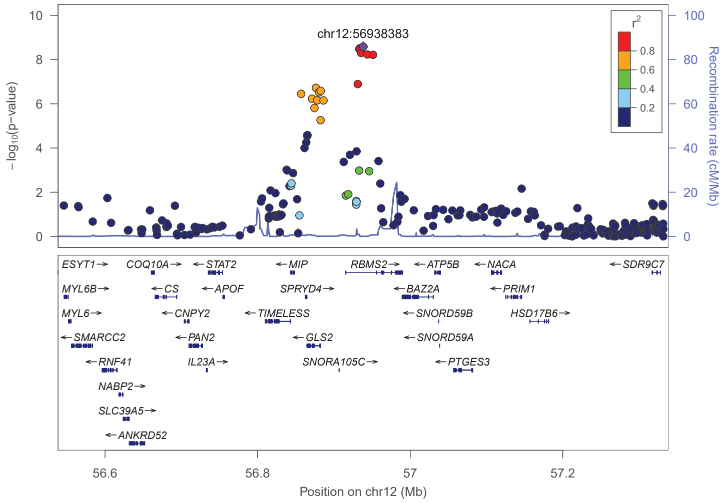
chr12:56938383
r2
0.8
0.6
0.4
0.2
−log10(p−value)
Recombination rate (cM/Mb)
ESYT1→
COQ10A→
←STAT2
←MIP
RBMS2→
←ATP5B
←NACA
←SDR9C7
MYL6B→
←CS
←APOF
SPRYD4→
←BAZ2A
←PRIM1
MYL6→
←CNPY2
←TIMELESS
←SNORD59B
HSD17B6→
←SMARCC2
←PAN2
←GLS2
←SNORD59A
←RNF41
IL23A→
SNORA105C→
←PTGES3
NABP2→
SLC39A5→
←ANKRD52
56.6
56.8
57
57.2
Position on chr12 (Mb)

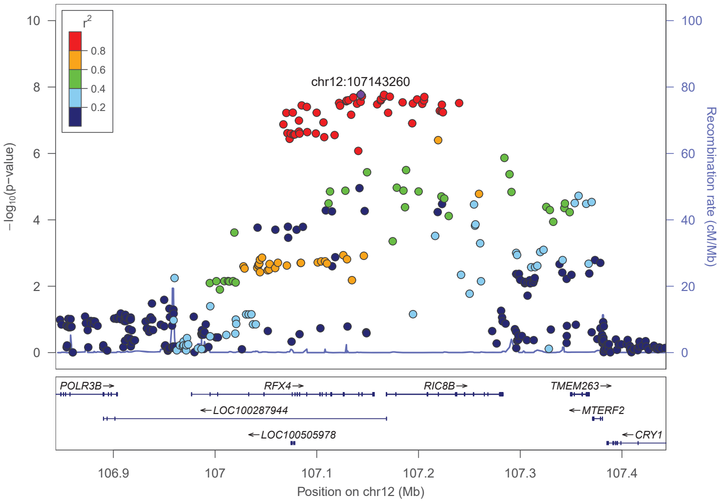
chr12:107143260
r2
0.8
0.6
0.4
0.2
−log10(p−value)
Recombination rate (cM/Mb)
POLR3B→
RFX4→
RIC8B→
TMEM263→
←LOC100287944
←MTERF2
←LOC100505978
←CRY1
106.9
107
107.1
107.2
107.3
107.4
Position on chr12 (Mb)

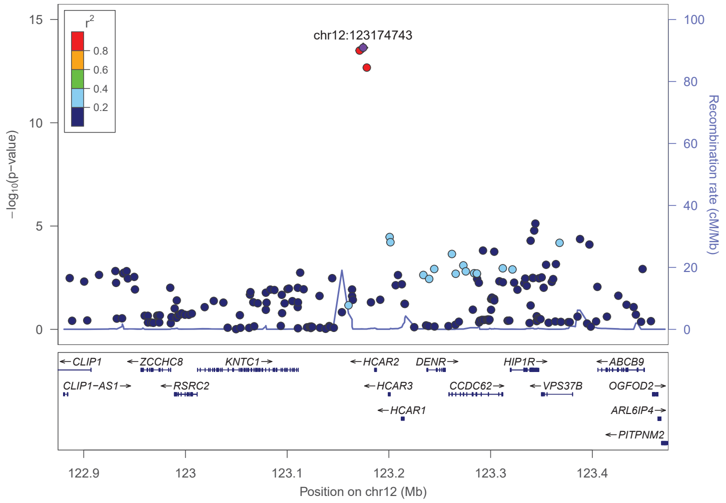
chr12:123174743
r2
0.8
0.6
0.4
0.2
−log10(p−value)
Recombination rate (cM/Mb)
←CLIP1
←ZCCHC8
KNTC1→
←HCAR2
DENR→
HIP1R→
←ABCB9
CLIP1−AS1→
←RSRC2
←HCAR3
CCDC62→
←VPS37B
OGFOD2→
←HCAR1
ARL6IP4→
←PITPNM2
122.9
123
123.1
123.2
123.3
123.4
Position on chr12 (Mb)

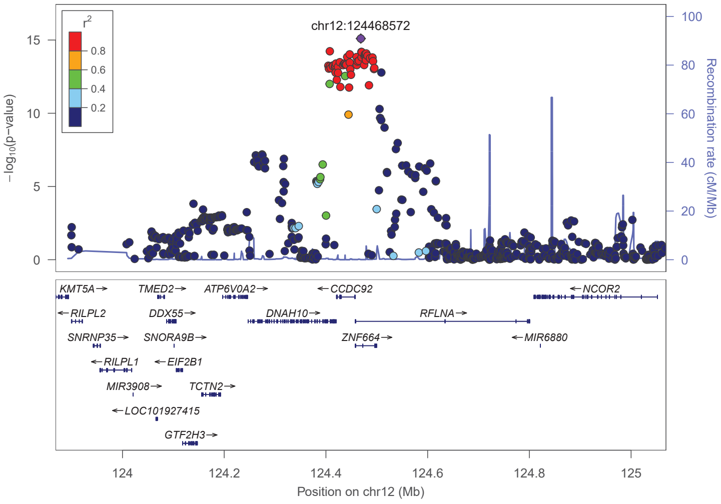
chr12:124468572
r2
0.8
0.6
0.4
0.2
−log10(p−value)
Recombination rate (cM/Mb)
KMT5A→
TMED2→
ATP6V0A2→
←CCDC92
←NCOR2
←RILPL2
DDX55→
DNAH10→
RFLNA→
SNRNP35→
SNORA9B→
ZNF664→
←MIR6880
←RILPL1
←EIF2B1
MIR3908→
TCTN2→
←LOC101927415
GTF2H3→
124
124.2
124.4
124.6
124.8
125
Position on chr12 (Mb)

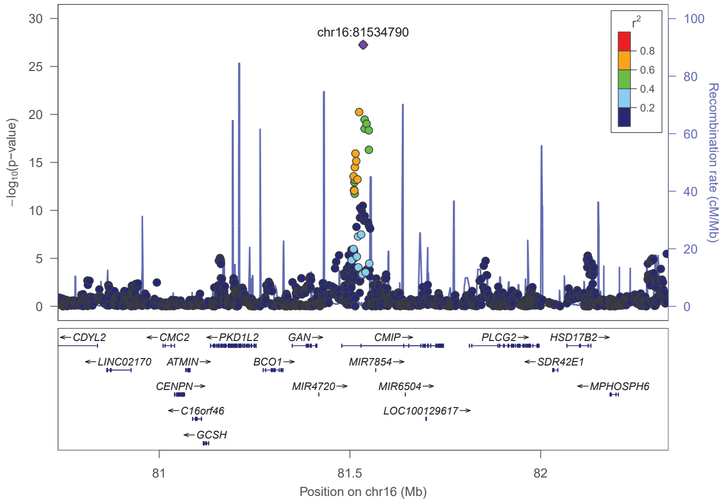
chr16:81534790
r2
0.8
0.6
0.4
0.2
−log10(p−value)
Recombination rate (cM/Mb)
←CDYL2
←CMC2
←PKD1L2
GAN→
CMIP→
PLCG2→
HSD17B2→
←LINC02170
ATMIN→
BCO1→
MIR7854→
←SDR42E1
CENPN→
MIR4720→
MIR6504→
←MPHOSPH6
←C16orf46
LOC100129617→
←GCSH
81
81.5
82
Position on chr16 (Mb)

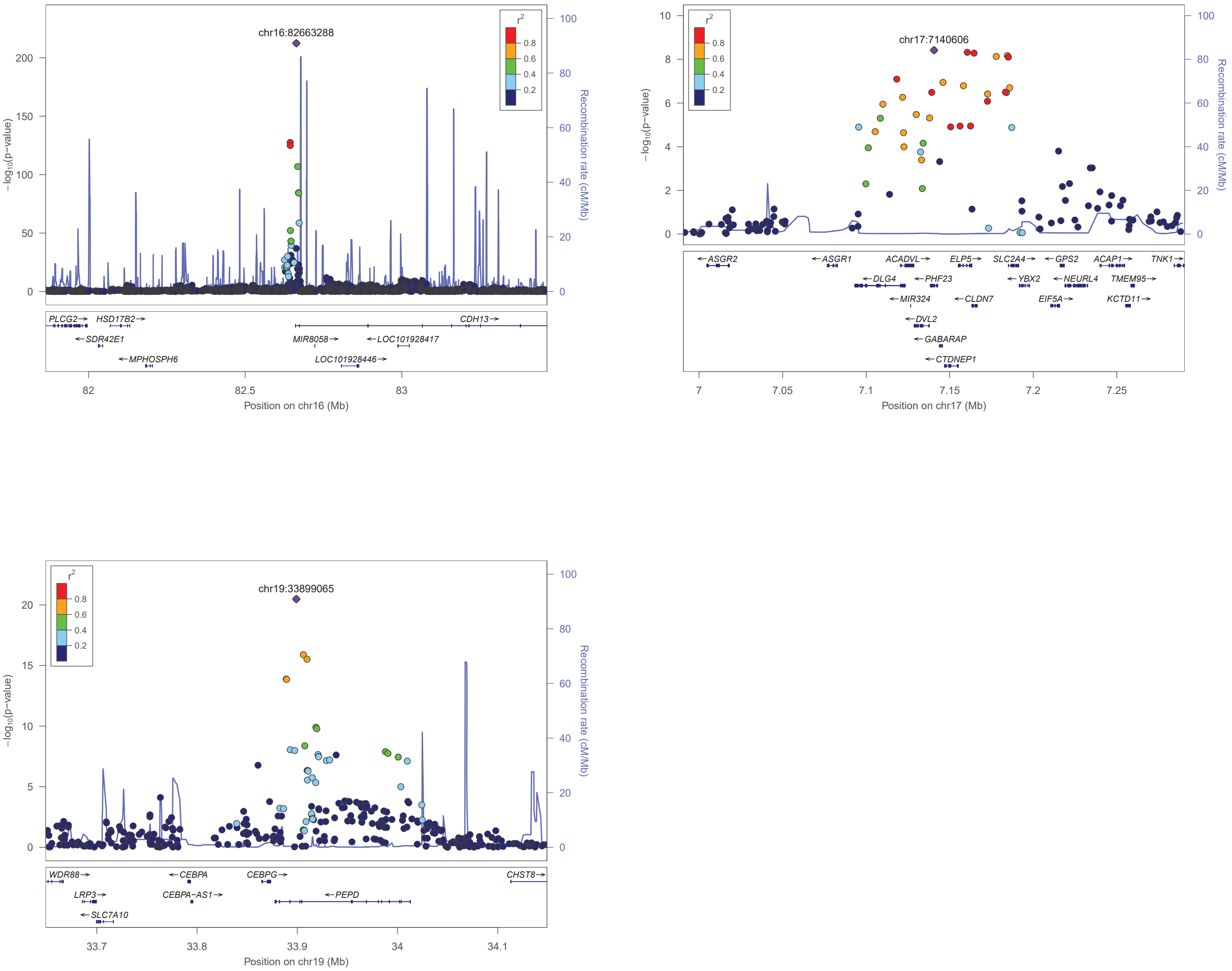


**Figure S1. Locus Zoom plots of multi-ancestry meta-analysis results**. Locus zoom plots of multi-ancestry summary statistics colored by LD to the lead variant. The loci are located at or near *CSF1, LYPAL1-AS1, IRS1, PBMR1, ADIPOQ, FAM13A, ARL15, VEGFA, RGS17, TRIB1, ADRB1, WDR11-FGFR2, PDE3B, PDE3A, RBMS2, RFX4, HCAR1, ZNF664, CMIP, CDH13, PHF23,* and *PEPD.*

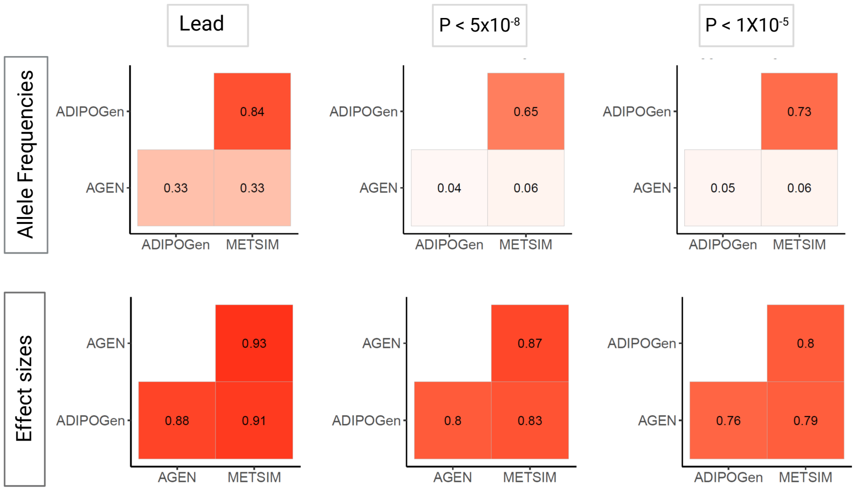


**Figure S2. Spearman correlation of effect sizes and allele frequencies between ADIPOGen, AGEN, and METSIM.** The top row of plots shows the correlation (r) values for the allele frequencies, and the bottom row of plots shows the correlation (r) values for the effect sizes. Each column represents a set of variants based on different p-value thresholds: the lead variant at a locus, all variants with P < $5 \times 10^{-8}$, and all variants with P < $1 \times 10^{-5}$. The boldness of the color is based on the strength of the r value, with darker colors indicating stronger correlations..

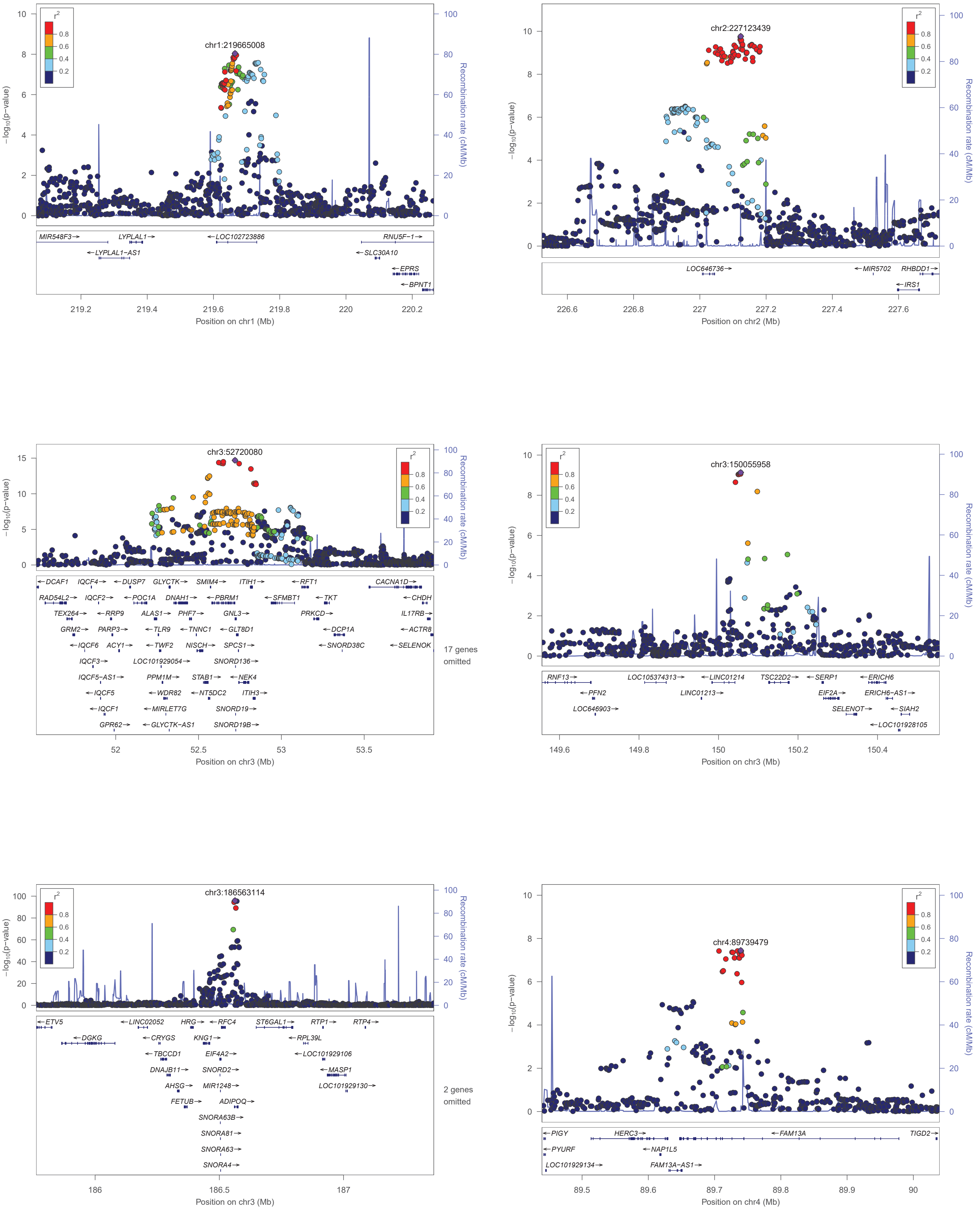

chr1:219665008
−log10(p−value)
Recombination rate (cM/Mb)
r2
0.8
0.6
0.4
0.2
MIR548F3→
LYPLAL1→
←LOC102723886
RNU5F−1→
←LYPLAL1−AS1
←SLC30A10
←EPRS
←BPNT1
219.2
219.4
219.6
219.8
220
220.2
Position on chr1 (Mb)
chr2:227123439
LOC646736→
←MIR5702
RHBDD1→
←IRS1
226.6
226.8
227
227.2
227.4
227.6
Position on chr2 (Mb)
chr3:52720080
←DCAF1
IQCF4→
←DUSP7
GLYCTK→
SMIM4→
ITIH1→
←RFT1
CACNA1D→
RAD54L2→
IQCF2→
←POC1A
DNAH1→
←PBRM1
←SFMBT1
←TKT
←CHDH
TEX264→
←RRP9
ALAS1→
PHF7→
GNL3→
PRKCD→
IL17RB→
GRM2→
PARP3→
←TLR9
←TNNC1
←GLT8D1
←DCP1A
←ACTR8
←IQCF6
ACY1→
←TWF2
NISCH→
SPCS1→
←SNORD38C
←SELENOK
IQCF3→
LOC101929054→
SNORD136→
IQCF5−AS1→
PPM1M→
STAB1→
←NEK4
←IQCF5
←WDR82
←NT5DC2
ITIH3→
←IQCF1
←MIRLET7G
SNORD19→
GPR62→
←GLYCTK−AS1
SNORD19B→
17 genes omitted
52
52.5
53
53.5
Position on chr3 (Mb)
chr3:150055958
RNF13→
LOC105374313→
←LINC01214
TSC22D2→
←SERP1
←ERICH6
←PFN2
LINC01213→
EIF2A→
ERICH6−AS1→
LOC646903→
SELENOT→
←SIAH2
←LOC101928105
149.6
149.8
150
150.2
150.4
Position on chr3 (Mb)
chr3:186563114
←ETV5
←LINC02052
HRG→
←RFC4
ST6GAL1→
RTP1→
RTP4→
←DGKG
←CRYGS
KNG1→
←RPL39L
←TBCCD1
EIF4A2→
←LOC101929106
DNAJB11→
SNORD2→
←MASP1
AHSG→
MIR1248→
LOC101929130→
FETUB→
ADIPOQ→
SNORA63B→
SNORA81→
SNORA63→
SNORA4→
2 genes omitted
186
186.5
187
Position on chr3 (Mb)
chr4:89739479
←PIGY
HERC3→
←FAM13A
TIGD2→
←PYURF
←NAP1L5
LOC101929134→
FAM13A−AS1→
89.5
89.6
89.7
89.8
89.9
90
Position on chr4 (Mb)

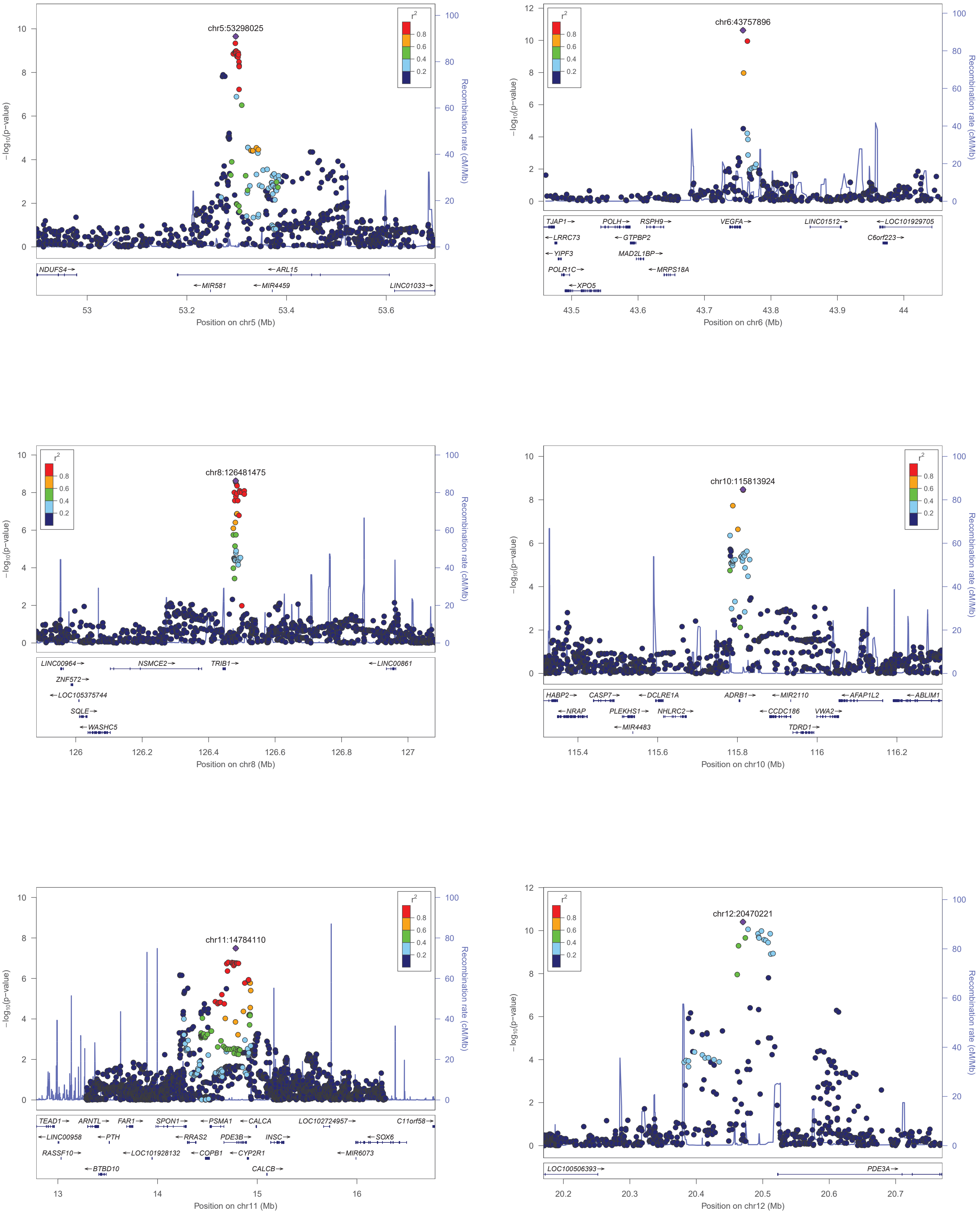
chr5:53298025
−log10(p−value)
Recombination rate (cM/Mb)
r2
NDUFS4→
←ARL15
←MIR581
←MIR4459
LINC01033→
Position on chr5 (Mb)
chr6:43757896
TJAP1→
POLH→
RSPH9→
VEGFA→
LINC01512→
←LOC101929705
←LRRC73
←GTPBP2
C6orf223→
←YIPF3
MAD2L1BP→
POLR1C→
←MRPS18A
←XPO5
Position on chr6 (Mb)
chr8:126481475
LINC00964→
NSMCE2→
TRIB1→
←LINC00861
ZNF572→
←LOC105375744
SQLE→
←WASHC5
Position on chr8 (Mb)
chr10:115813924
HABP2→
CASP7→
←DCLRE1A
ADRB1→
←MIR2110
←AFAP1L2
←ABLIM1
←NRAP
PLEKHS1→
NHLRC2→
←CCDC186
VWA2→
←MIR4483
TDRD1→
Position on chr10 (Mb)
chr11:14784110
TEAD1→
ARNTL→
FAR1→
SPON1→
←PSMA1
←CALCA
LOC102724957→
C11orf58→
←LINC00958
←PTH
←RRAS2
PDE3B→
INSC→
←SOX6
RASSF10→
←LOC101928132
←COPB1
←CYP2R1
←MIR6073
←BTBD10
CALCB→
Position on chr11 (Mb)
chr12:20470221
LOC100506393→
PDE3A→
Position on chr12 (Mb)

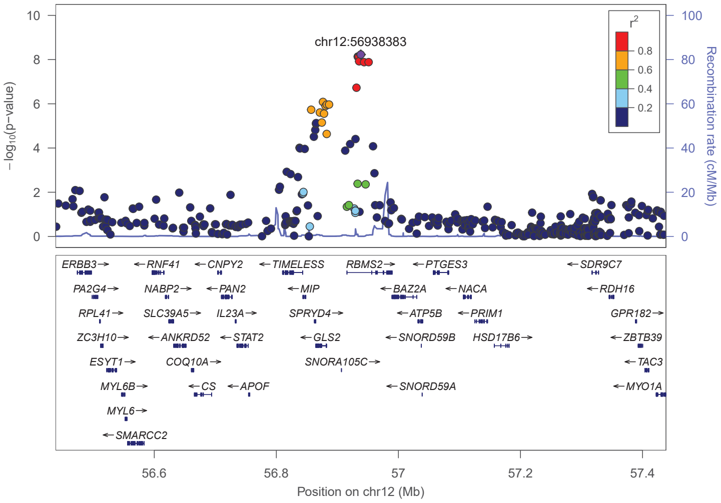
chr12:56938383
−log10(p−value)
Recombination rate (cM/Mb)
r2
ERBB3 RNF41 CNPY2 TIMELESS RBMS2 PTGES3 SDR9C7
PA2G4 NABP2 PAN2 MIP BAZ2A NACA RDH16
RPL41 SLC39A5 IL23A SPRYD4 ATP5B PRIM1 GPR182
ZC3H10 ANKRD52 STAT2 GLS2 SNORD59B HSD17B6 ZBTB39
ESYT1 COQ10A SNORA105C TAC3
MYL6B CS APOF SNORD59A MYO1A
MYL6
SMARCC2
Position on chr12 (Mb)

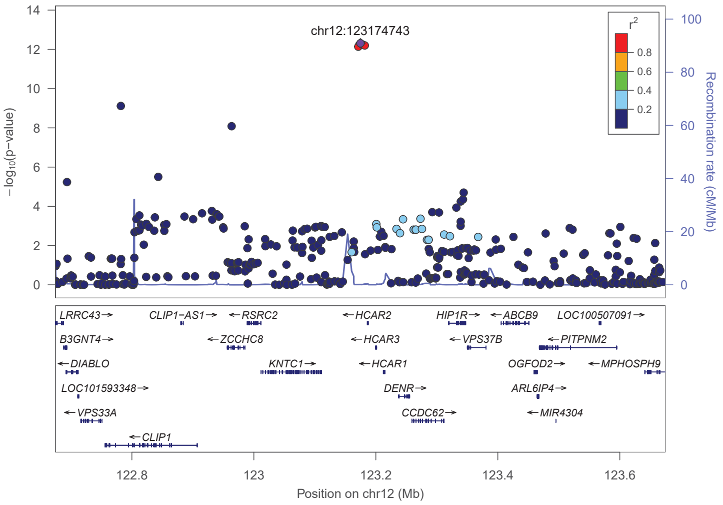
chr12:123174743
−log10(p−value)
Recombination rate (cM/Mb)
r2
LRRC43 CLIP1−AS1 RSRC2 HCAR2 HIP1R ABCB9 LOC100507091
B3GNT4 ZCCHC8 HCAR3 VPS37B PITPNM2
DIABLO KNTC1 HCAR1 OGFOD2 MPHOSPH9
LOC101593348 DENR ARL6IP4
VPS33A CCDC62 MIR4304
CLIP1
Position on chr12 (Mb)

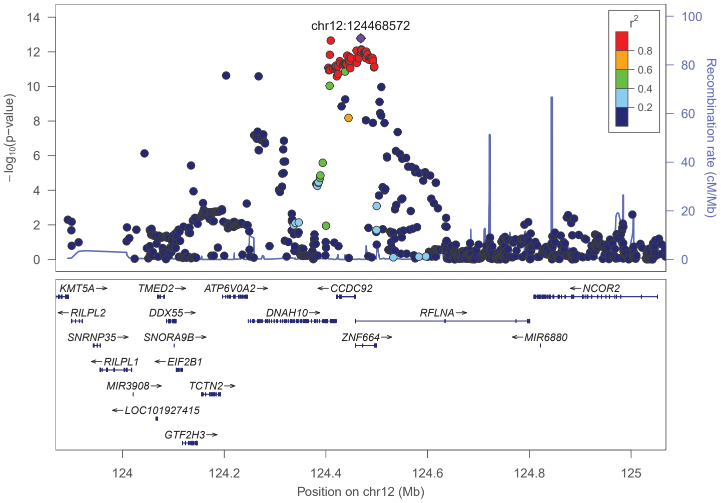
chr12:124468572
−log10(p−value)
Recombination rate (cM/Mb)
r2
KMT5A TMED2 ATP6V0A2 CCDC92 NCOR2
RILPL2 DDX55 DNAH10 RFLNA
SNRNP35 SNORA9B ZNF664 MIR6880
RILPL1 EIF2B1
MIR3908 TCTN2
LOC101927415
GTF2H3
Position on chr12 (Mb)

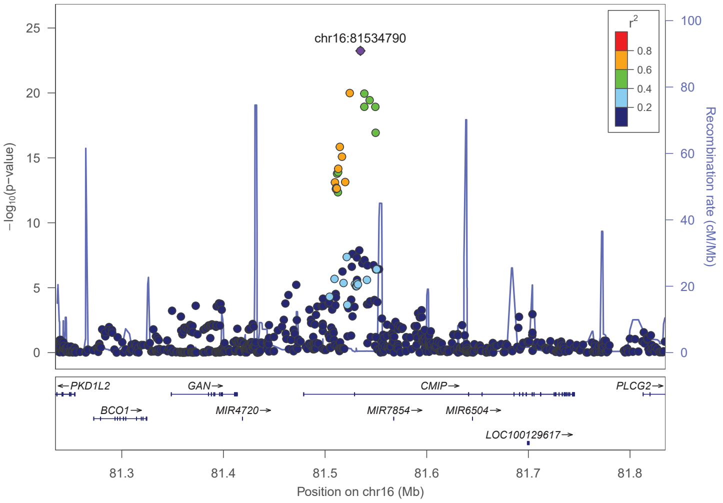
chr16:81534790
−log10(p−value)
Recombination rate (cM/Mb)
r2
PKD1L2 GAN CMIP PLCG2
BCO1 MIR4720 MIR7854 MIR6504
LOC100129617
Position on chr16 (Mb)

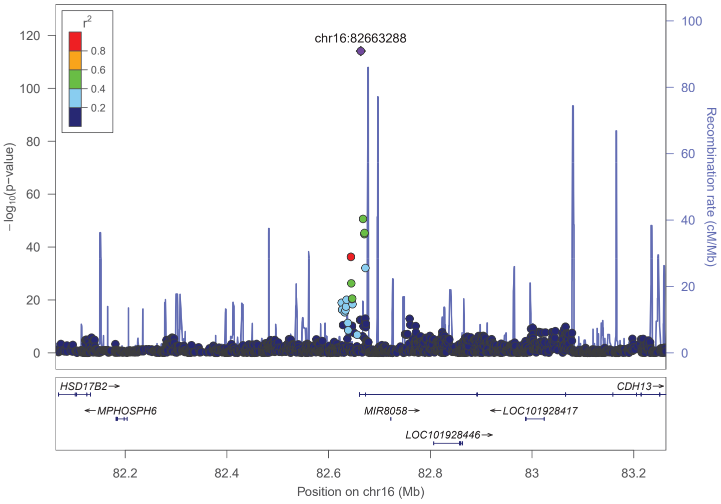
chr16:82663288
−log10(p−value)
Recombination rate (cM/Mb)
r2
HSD17B2 CDH13
MPHOSPH6 MIR8058 LOC101928417
LOC101928446
Position on chr16 (Mb)

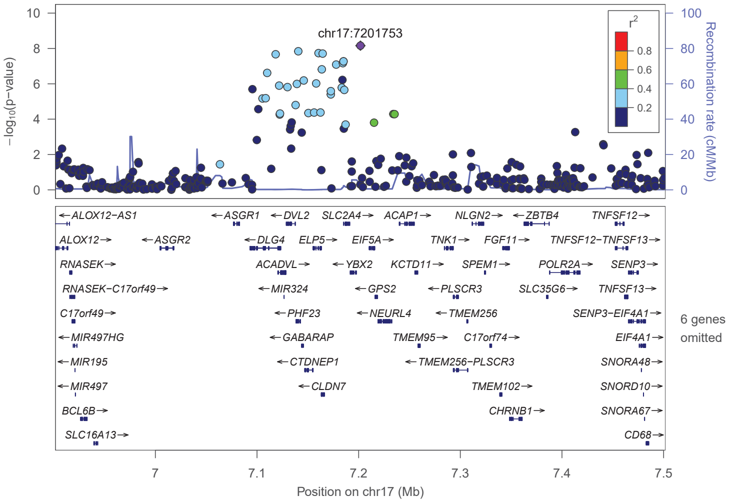
chr17:7201753
−log10(p−value)
Recombination rate (cM/Mb)
r2
ALOX12−AS1 ASGR1 DVL2 SLC2A4 ACAP1 NLGN2 ZBTB4 TNFSF12
ALOX12 ASGR2 DLG4 ELP5 EIF5A TNK1 FGF11 TNFSF12−TNFSF13
RNASEK ACADVL YBX2 KCTD11 SPEM1 POLR2A SENP3
RNASEK−C17orf49 MIR324 GPS2 PLSCR3 SLC35G6 TNFSF13
C17orf49 PHF23 NEURL4 TMEM256 SENP3−EIF4A1
MIR497HG GABARAP TMEM95 C17orf74 EIF4A1
MIR195 CTDNEP1 TMEM256−PLSCR3 SNORA48
MIR497 CLDN7 TMEM102 SNORD10
BCL6B CHRNB1 SNORA67
SLC16A13 CD68
6 genes omitted
Position on chr17 (Mb)

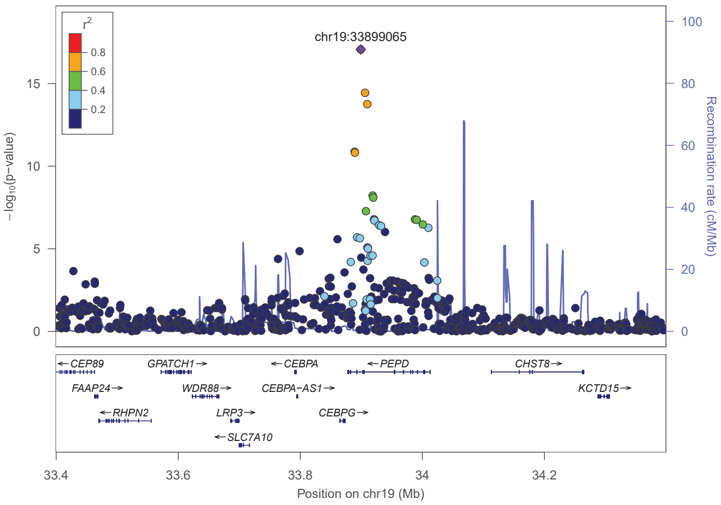


**Figure S3. Locus Zoom plots of European-ancestry meta-analysis results.** Locus zoom plots of European-ancestry summary results colored by LD to the lead variant. The loci are located at or near *LYPAL1-AS1, IRS1, PBMR1, LINC01214, ADIPOQ, FAM13A, ARL15, VEGFA, RGS17, TRIB1, ADRB1, PDE3B, PDE3A, RBMS2, YBX2, HCAR1, ZNF664, CMIP, CDH13, PHF23,* and *PEPD.*

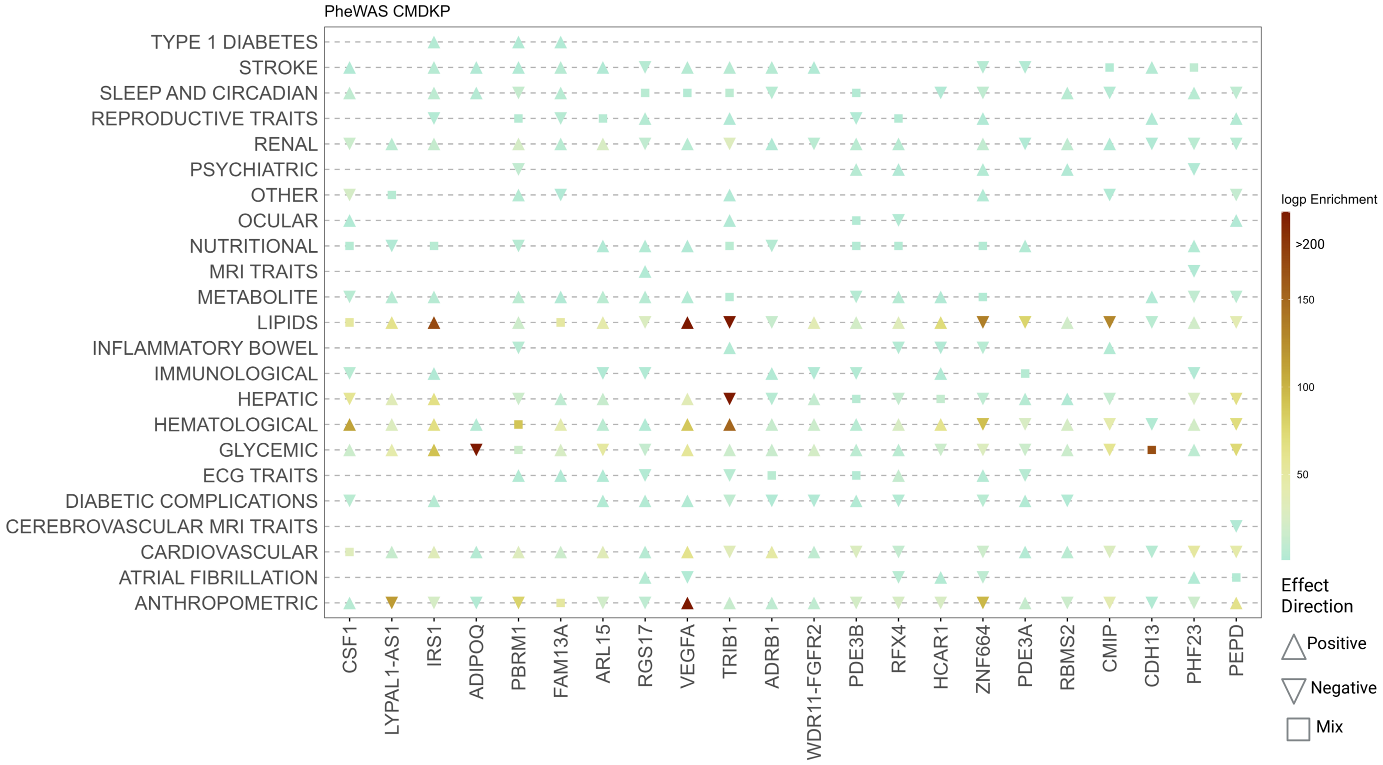


**Figure S4. Lead variants at multi-ancestry adiponectin-associated loci are pleiotropic** The Y-axis lists all phenotype groups from the Common Metabolic Diseases Knowledge Portal (CMDKP), many of which are comprised of multiple related traits. For each lead variant (denoted by the nearest gene to the lead variant on the x-axis), a triangle indicates at least one genome-wide significant association was previously reported for at least one trait within the phenotype group. The adiponectin-increasing allele was used as the effect allele. The direction of the triangle corresponds to the direction of effect for the phenotype(s) from the CMDKP: up indicates increased trait/disease risk, down indicates decreased trait/disease risk, and a square indicates traits within the phenotype group include a mix of both increased and decreased trait/disease risk. Points are colored based on the $-log_{10}$(P) value of the strongest previously reported association.

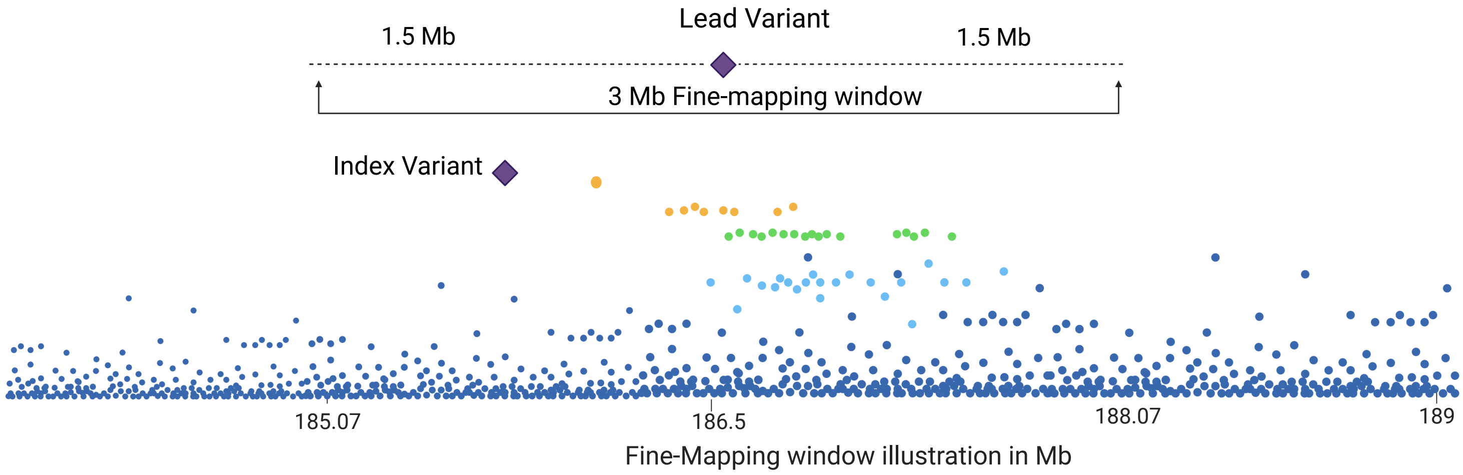


**Figure S5. Illustration of fine-mapping window** The "lead" genetic variants at each locus were defined as the most significant variant (P < $5 \times 10^{-8}$) within a 500 kb window. The fine-mapping window is the 3 Mb region (+/- 1.5 Mb) around the lead variant of each locus. In the European-ancestry meta-analysis only, we use "index variant" to represent distinct association signals identified by GCTA-COJO at a locus if 1) the variant achieved P < $5 \times 10^{-8}$ in the GCTA cojo analysis, 2) the variant was located within +/- 1 Mb from the original lead variant at that locus, and 3) the variant had an initial P-value in the meta-analysis of P<0.05.

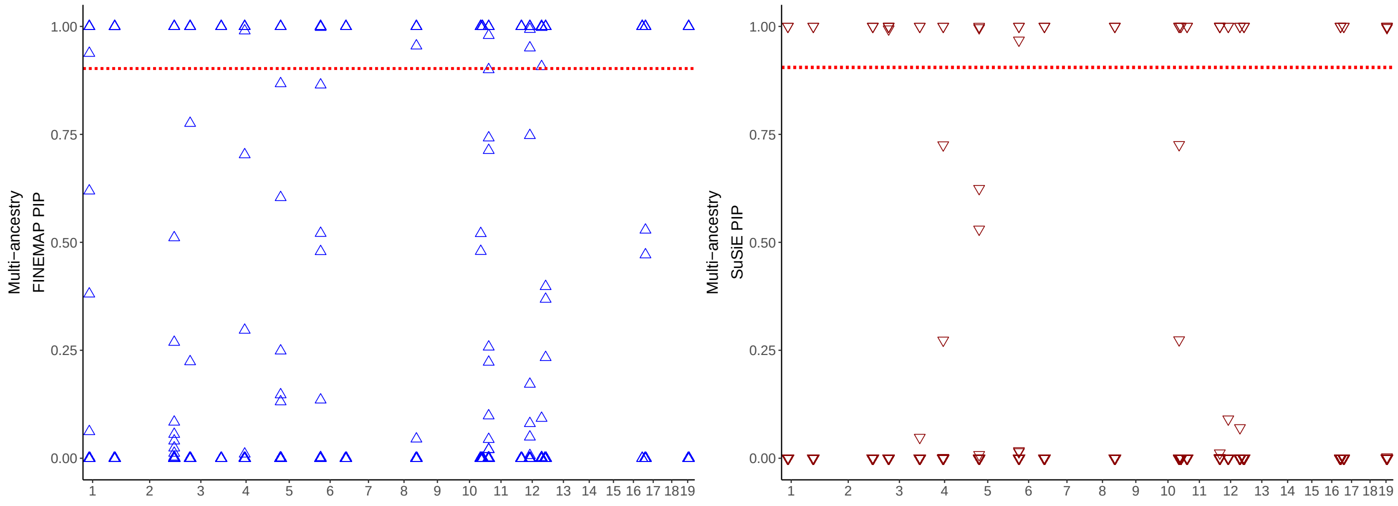


**Figure S6. Overview of fine-mapped variants of multi-ancestry meta-analysis using FINEMAP+SuSiE.** The y-axis denotes the posterior probabilities (PIP) reported by FINEMAP (left) or SuSiE (right). The red dotted line indicates the PIP $> 0.90$.

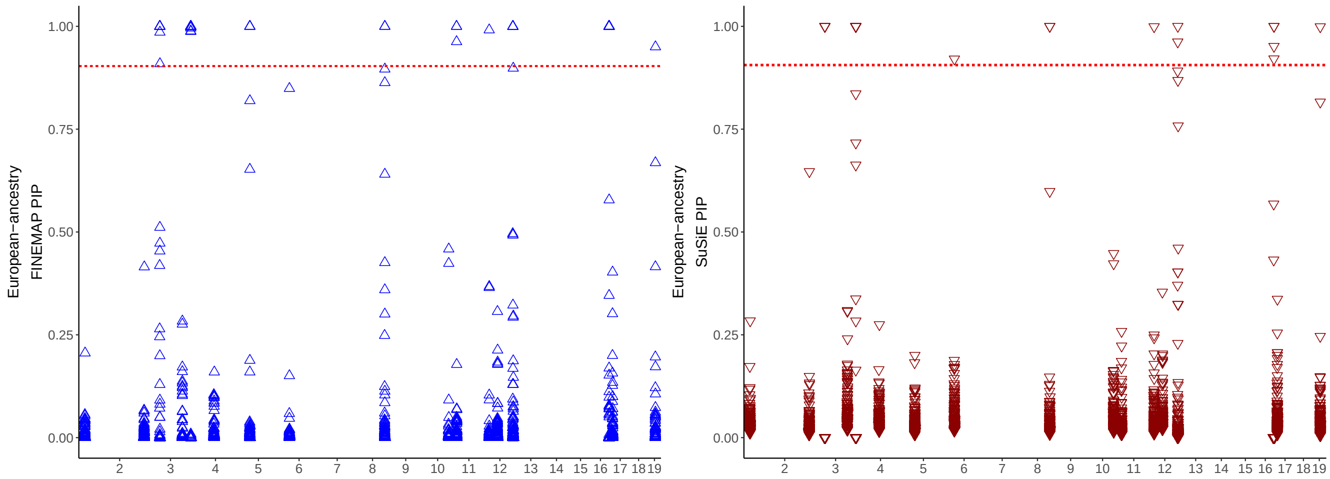


**Figure S7. Overview of fine-mapped variants of European-ancestry meta-analysis using FINEMAP+SuSiE.** Thy y-axis denotes the posterior probabilities (PIP) reported by FINEMAP (left) or SuSiE (right). The red dotted line indicates the PIP $> 0.90$.

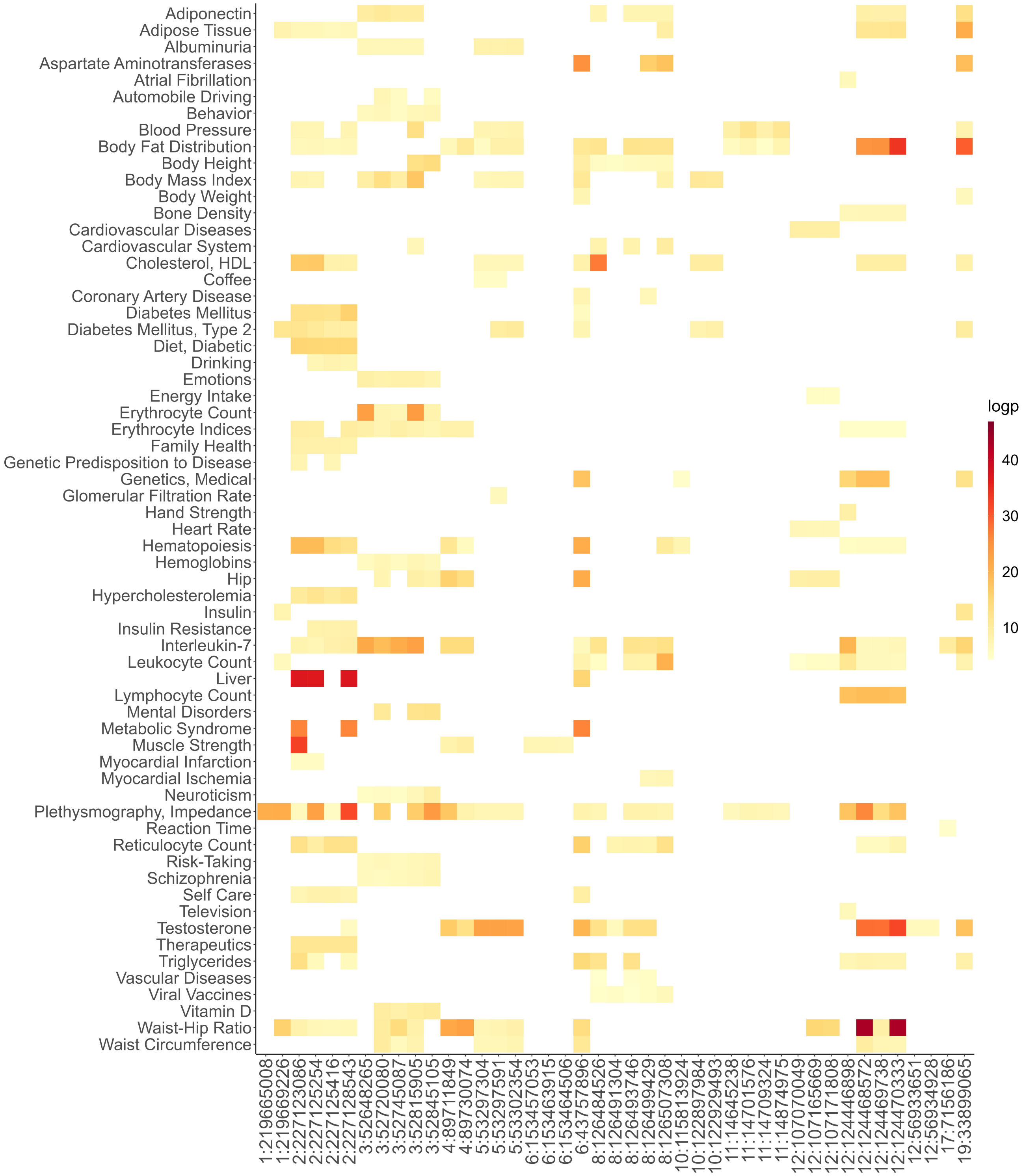


**Figure S8. Overview of pleiotropy of nominated adiponectin causal variants.** The enrichment of nominated causal variants denoted by (log P) value with 2,629 traits from the CAUSALdb database. The x-axis represents the chromosome and position of the nominated causal variant.

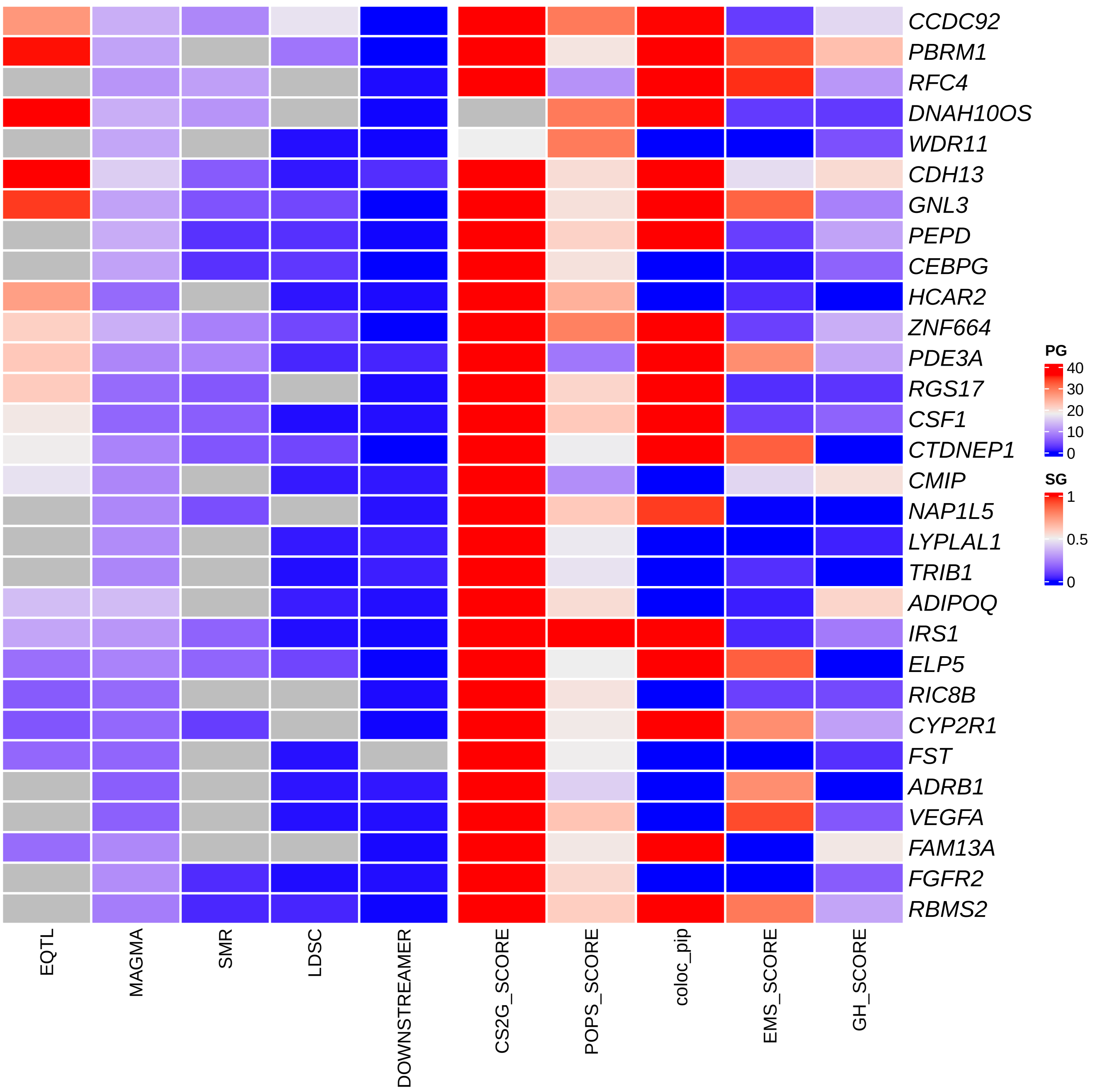


**Figure S9. Values of gene prioritization strategies for 30 effector genes in a multi-ancestry analysis.** The values of gene prioritization strategies are either shown in $-log10(p)$ enrichment or a scaled score of 1-10, depending on the strategy for the 30 prioritized genes.

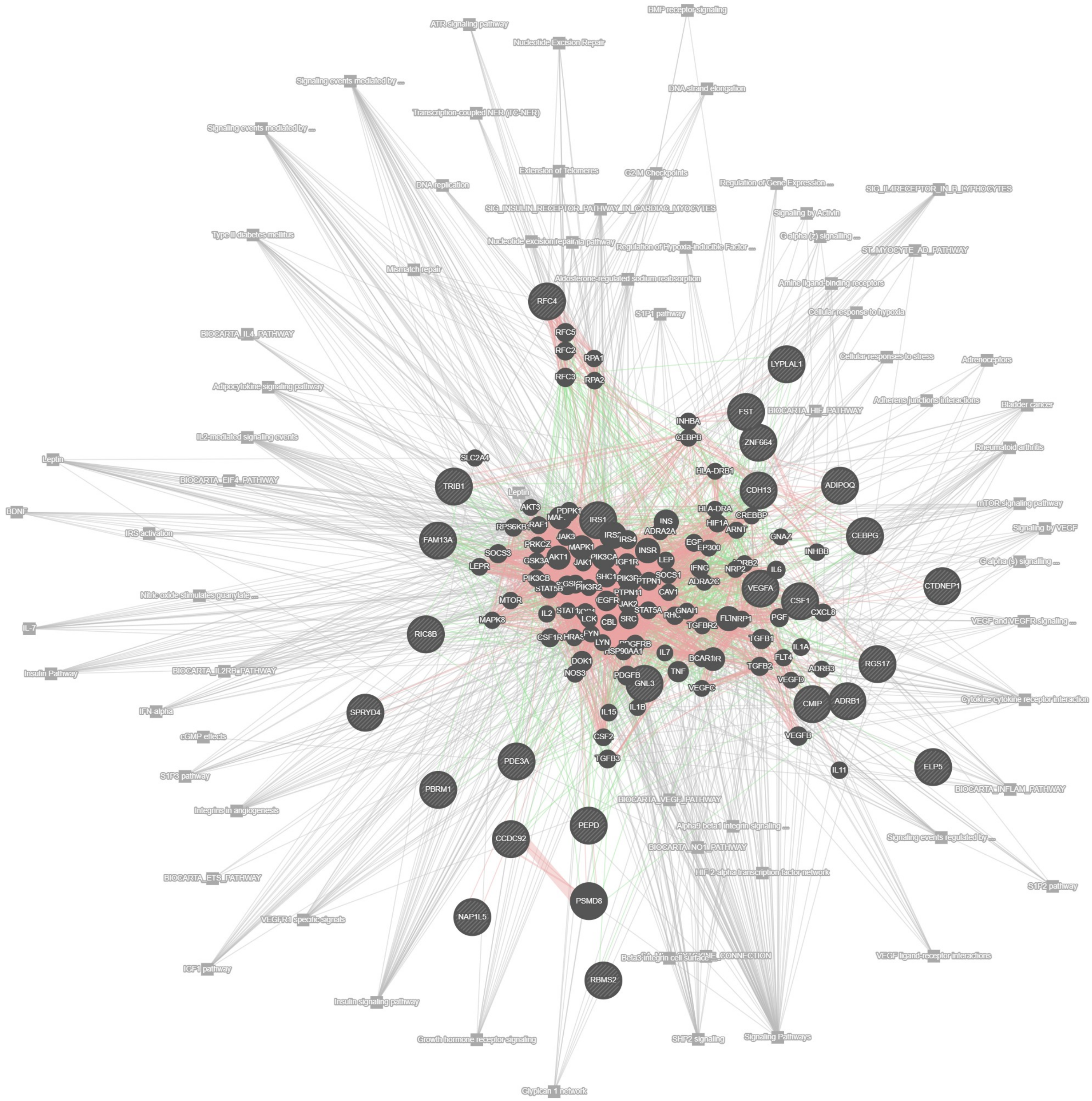


**Figure S10. Full functional interaction network of adiponectin multi-ancestry prioritized genes.** A complete interactive functional association network, illustrating the relationships among the connected genes, prioritized genes, and underlying associated pathways constructed using GeneMANIA.
